# A decision analytic framework for triggering cholera outbreak response based on early-case surveillance

**DOI:** 10.64898/2026.07.16.26358045

**Authors:** Christina Alam, Qulu Zheng, Javier Perez-Saez, Jong-Hoon Kim, Andrew S. Azman, Elizabeth C. Lee

## Abstract

**Background:** Cholera outbreaks can spread rapidly, which means that the optimal decision-making window for a large, coordinated response is very narrow. Alerts for triggering interventions need to balance tradeoffs between wasting resources on false positives and delaying decisions until they lose effectiveness. A systematic evaluation of such tradeoffs across settings is needed to understand which alerts may have the greatest public health utility and where.

**Methods:** Using weekly suspected cholera surveillance across 4,081 subnational administrative units across 34 countries in Africa from 2010-2023, we evaluated 24 alert definitions (4 alert types with different numeric thresholds) over a 1-year post-alert period on five utility dimensions – potential health impact, potential intervention efficiency, positive predictive value (PPV) for large outbreaks, proportion of missed outbreaks, and timeliness of alert trigger. The dimensions were combined into a utility score, which was used to identify the best alert across the continent and by country. For top-performing alerts, we estimated the reduction in potential health impact for additional delays in response using Bayesian hierarchical models.

**Results:** Fifty suspected cases for three consecutive weeks was the definition with the highest utility score across most contexts. In administrative units with 50,000 to 500,000 people, the year following such an alert experienced a mean of 376 suspected cases (standard deviation: 618.3) and 2 cases per 1000 population (SD: 4.1). Forty percent of such alerts (N alerts: 265) were followed by a 1-year period with over 300 cases, yet the definition missed 40% of outbreaks with over 300 cases (N outbreaks: 278) and was triggered 5.3 weeks (SD: 5) after outbreak start. Each week of delay was estimated to result in an additional 20% reduction (95% CrI: −23 to −17) of potential health impact in the outbreak response. One hundred cases over a three-week period was another definition that had high utility, particularly in administrative units with smaller populations and country-specific evaluations.

**Conclusion:** We present a decision analytic framework that can be deployed in a short decision-making window using case-based surveillance to trigger large-scale cholera response activities with moderately high utility across most African transmission contexts. Future work should consider adaptations based on local data availability and priorities and examine the generalizability of early case-based signals outside Africa.

## Introduction

Cholera is a waterborne bacterial disease that remains a significant public health threat in regions where the population lacks access to safe drinking water and adequate sanitation. While areas with endemic cholera transmission are unquestionably a high priority for continually-enhanced surveillance, prevention, and response, roughly 633 million people in Africa live under the significant uncertainty of sporadic cholera outbreaks [1]. Experiences in Malawi, Zambia, and Ghana from 2022 to 2024 demonstrated that even areas with extended periods of low activity could still be vulnerable to devastating cholera outbreaks that overwhelm health systems and spread to neighboring areas [1–4]. As such, surveillance officials in outbreak-prone areas are challenged with deciding when to raise the broader alarm; since cholera treatment facilities do not operate continuously in these locations, mortality may be particularly high when large outbreaks do indeed occur.

The rapidly explosive nature of cholera outbreaks and the relatively short average duration (three to five months) [5,6] motivate the need for timely coordination of large-scale responses like mass reactive oral cholera vaccination (OCV) campaigns and case-area targeted interventions (CATIs). Modeling studies suggested that initiating OCV campaigns within 15 days of outbreak onset could reduce cases and deaths by up to 80% [7], with impact declining by 24% for each week of delay [8]. Likewise, modeling showed that CATIs involving antibiotics and point-of-use water treatment are more effective when implemented before the outbreak transmission peak [9].

In reality, there is a narrow window during which decision-makers may have enough confidence to mount a large-scale response and for those actions to be early enough to significantly change the trajectory of an outbreak. Selection of early epidemic signals should maximize potential public health impact, predictive value, and timeliness, while also limiting costs associated with mobilizing resources in response to a false alarm. Existing frameworks for early outbreak identification have been explored in disease-agnostic settings and for meningococcal meningitis [10,11], but tradeoffs across different dimensions of utility should be pathogen- and context-specific and have not been systematically explored for cholera outbreaks.

While current cholera surveillance guidance from the Global Task Force on Cholera Control (GTFCC) proposes case-based thresholds to signal suspected, probable, or confirmed cholera outbreaks [12], no systematic evaluation of what follows such an alert, including the expected outbreak size and the relative utility of mounting a large-scale response, has been conducted. Previous work reported significant variability in the time between outbreak identification and submission of an OCV request to the emergency stockpile (range of 12 to 206 days) [13], underscoring the need to link early surveillance-based alerts to future outbreak outcomes in order to reduce delays in implementation and uncertainty in decision-making.

We examine 24 early cholera alert definitions (4 alert types with multiple threshold values) as potential heuristics for triggering large-scale interventions using a large collection of cholera incidence data from outbreak-prone African locations from 2010 to 2023. Multiple dimensions of early alert utility – potential health impact, potential intervention efficiency, positive predictive value (PPV) for large outbreaks, proportion of missed outbreaks, and timeliness of alert trigger – are combined into a decision analytic framework to evaluate alert definition performance. We assess tradeoffs between the utility dimensions and stringency of alert thresholds and identify alert definitions with high utility across different population sizes and countries. We then use Bayesian hierarchical models to estimate how delays in implementation may affect the potential health impact of an intervention.

## Methods

We applied 24 alert definitions consisting of short case-based surveillance signals to unified cholera incidence time series in outbreak-prone locations. We then investigated the 1-year period following the alerts, converting metrics into a multidimensional score that encompasses utility tradeoffs. Utility scores were used to assess the performance of alerts with different stringencies (threshold values) and identify the top-performing alert definitions in different population contexts. We then estimated the impact of implementation delays on potential health impact of interventions for top-performing definitions.

### Data collection and preparation

Weekly consecutive cholera incidence time series representing January 1, 2010 to December 31, 2023 for subnational administrative units in Africa (4,081 units in 34 countries) were prepared using data from a global cholera database [14] that includes suspected and bacteriologically confirmed cholera incidence data from public and non-public data sources [6] (Supp Fig. 1). In brief, daily cholera incidence data from locations at administrative levels 1, 2, and 3 were aggregated to weeks and merged with observations originally reported at weekly scale. Observations reporting data for the same location and week were averaged. Locations were audited for validity and linked to shapefiles in order to generate UN-adjusted annual population estimates from WorldPop [15]. As population estimates were linked to locations annually, there were a total of 4284 unique location and annual population size combinations (location-population combinations).

Locations with less than two weeks of data or missing shapefiles were removed from analysis. Locations with endemic cholera transmission (Supp Fig. 2), defined as those with at least 3 years of data and reporting non-zero suspected cholera cases for at least 50% of those weeks (Supp Fig. 3), were removed from analysis because the thresholds suitable for outbreak-prone locations were too low to also serve as meaningful alerts in endemic locations.

### Ethics

The Johns Hopkins Bloomberg School of Public Health (BSPH) Institutional Review Board (IRB) declared the collation and use of the cholera surveillance data to be exempt (BSPH IRB No. 27682).

### Alert definitions

We define alert definitions as short case-based patterns in cholera incidence time series with potential to indicate increasing cholera activity preceding a large outbreak. We considered alert definitions of four types with multiple threshold values: 1) trend-based alerts, which are triggered by an upward case trend of *n* weeks relative to the mean of the previous four weeks, 2) weekly case-based alerts, when at least *x* suspected cases are observed for three consecutive weeks, 3) cumulative case-based alerts, when at least *x* suspected cases are observed over a three-week period, and 4) weekly rate-based alerts, when at least *x* suspected cases per 10,000 people are observed for three consecutive weeks (Supp Fig. 4, Supp Fig. 5, Supp Methods).

Threshold values were ranked by stringency within an alert type. For example, within weekly case-based alerts, ≥ 2 weekly cases had the lowest stringency (1) while ≥ 250 weekly cases had the highest stringency (7). The three trend-based alerts were assigned intermediate stringency values (2, 4, and 6 for 1-week, 2-week, and 3-week trends, respectively) in order to remain on a comparable scale to the others. Due to the nested nature of the thresholds by alert type, lower stringency alerts are triggered earlier and more often than higher stringency alerts.

We also explored the value of layering confirmed cholera case thresholds to lower stringency alert definitions for the subset of alerts reporting bacteriologically confirmed case data (Supp Methods, Supp Fig. 6).

### Multidimensional utility framework

The 24 alert definitions were evaluated for their utility across five quantitative dimensions: 1) potential impact: number of suspected incident cases in the evaluation period, 2) potential efficiency: impact standardized by population size, 3) positive predictive value (PPV): proportion of alerts with over 300 suspected cases in the evaluation period, 4) missed outbreaks: proportion of outbreaks over 300 cases missed, and 5) timeliness: timeliness of the alert relative to the outbreak start (Table 1, Supp Methods).

**Table 1.** Dimensions of alert definition utility as an early warning signal of large cholera outbreaks. The table describes each dimension, the motivation for including it in a utility score, and a description of how it was translated into a quantitative metric.

| Alert utility dimension | Motivation | Quantitative metric |
| --- | --- | --- |
| Potential Impact | A useful alert helps to maximize the public health impact of outbreak control measures like reactive OCV campaigns | Mean suspected cases observed in the 52-week evaluation period after alert. |
| Potential Efficiency | A useful alert helps to maximize the impact per target population (here, per dose of reactive OCV | Mean suspected cases per target population in the 52-week evaluation period after alert. |
|  | campaigns). |  |
| Positive Predictive Value (PPV) | A useful alert has a high probability of being followed by a large number of cases. | Proportion of alerts preceding at least 300 suspected cases in the 52-week evaluation period after alert. |
| Missed Outbreaks | A useful alert minimizes the proportion of large outbreaks missed. | Proportion of outbreaks of at least 300 suspected cases missed by the alert. |
| Timeliness | A useful alert is triggered as early in a large outbreak as possible. | Mean lag in weeks between outbreak start and alert trigger for outbreaks of at least 300 suspected cases |

The impact, efficiency, and PPV metrics were calculated for a 1-year post-alert evaluation period that begins following an alert and “implementation period” when we assume control activities are being mounted. An implementation period of 4 weeks was selected for the primary analyses. Alerts with a partially-censored evaluation period due to missingness were assumed to have zero reported cases during these weeks, similar to previous studies [6].

The missed and timeliness metrics were calculated by linking alerts to the occurrence of “cholera outbreaks,” thus allowing us to identify “false negative” signals and evaluate the timeliness of alert triggers (Supp Methods, Supp Fig. 7). We applied a systematic, previously-described outbreak definition [6,16] to the same cholera surveillance dataset and considered only outbreaks that exceeded 300 suspected cases. Timeliness values could be negative if an alert was triggered prior to an outbreak start.

Metrics were centered and standardized to calculate a dimension score; where appropriate, dimension scores were multiplied by −1 so that larger positive values of the score represented higher utility (missed, timeliness). The overall alert utility score was calculated as the sum of the five dimension scores. Top-performing alert definitions were identified by ranking the utility scores of alert definitions that had no dimension scores less than or equal to −1. Utility scores and top-performing alert definitions were identified for multiple “population contexts” – by population group for the entire continent (<50k, 50-500k, and >500k people) and by population group for each country. Alternative dimension thresholds and a rank-sum-based utility score were considered in sensitivity analyses (Supp Tab. 1-3, Supp Fig. 8-9).

### Public access

We prepared an R Shiny web application that enables users to set expectations about different utility dimensions following an alert. The application displays summary statistics and distributions of dimension metrics for different population contexts for all locations in Africa and by country. It also visualizes a random selection of cholera surveillance time series centered around an alert in order to provide insights about the shape of epidemic curves and speed of outbreaks following an alert. The application codebase is available at: https://github.com/GenevaIDD/cholera-alert-explorer-dashboard

### Estimating changes to impact after implementation delay

We used Bayesian hierarchical regression models to estimate the effect of delay on alert impact by modeling the ratio of cases in the evaluation period after a delay of *x* weeks (where *x* was 1 to 24 in weekly intervals) to that after a delay of 0 weeks. Alert definition and continent-level population group combinations were modeled separately.

Random intercepts were used to account for heterogeneity across scales and leverage partial pooling to estimate the effect of delays at levels with limited data. We compared seven model structures with different combinations of country, administrative unit level 1, location, and alert-specific effects (Supp Tab. 4). From these, we selected the parsimonious best-fit model after evaluating convergence traceplots, R-hat [17], Pareto smoothed importance-sampling leave-one-out cross-validation (PSIS-LOO) (Supp Tab. 5) [18], and the Watanabe-Akaike Information Criterion (WAIC) (Supp Tab. 6) from the loo R package (Supp Methods) [19]. Models were fit with Stan’s implementation of Hamiltonian Monte Carlo via the rstan package in R [20,21] with four parallel chains, each with 3000 warm up iterations and 3000 samples.

## Results

Of 4127 subnational locations in the dataset, 4081 (98.9%) were determined to be outbreak-prone locations (henceforth “locations”), and 46 (1.1%) were considered endemic and were removed from analysis (Supp Fig. 2). Among the 4284 location-population combinations, 533 (12.4%) had a population less than 50,000, 3123 (72.9%) had a population between 50,000 and 500,000, and 628 (14.7%) had a population over 500,000 (Supp Fig 1).

### Characterizing alert definitions in the utility framework

For locations with 50,000 to 500,000 people, which represents most of the available data, we first illustrate conceptual relationships between alert definition stringency, individual dimension scores, and the total utility score. Utility scores were highest among weekly case-, cumulative case- and rate-based alert definitions with moderate (4) to high (7) stringency (Fig 1A). Moderate to high stringency alert definitions were associated with higher impact, efficiency, and PPV, while low stringency alert definitions missed fewer outbreaks and were triggered after shorter delays relative to outbreak start (Fig 1B). Dimension scores had a monotonic relationship with stringency, except in instances where less than 30 alerts were triggered with high stringency definitions (possibly due to the small sample size). The impact, efficiency, and PPV dimension scores moved in the same direction, increasing with alert stringency. By contrast, the missed and timeliness dimensions scores moved in the same direction, decreasing with alert stringency (Supp Fig. 10, Supp Tab. 7).

**Figure 1.**
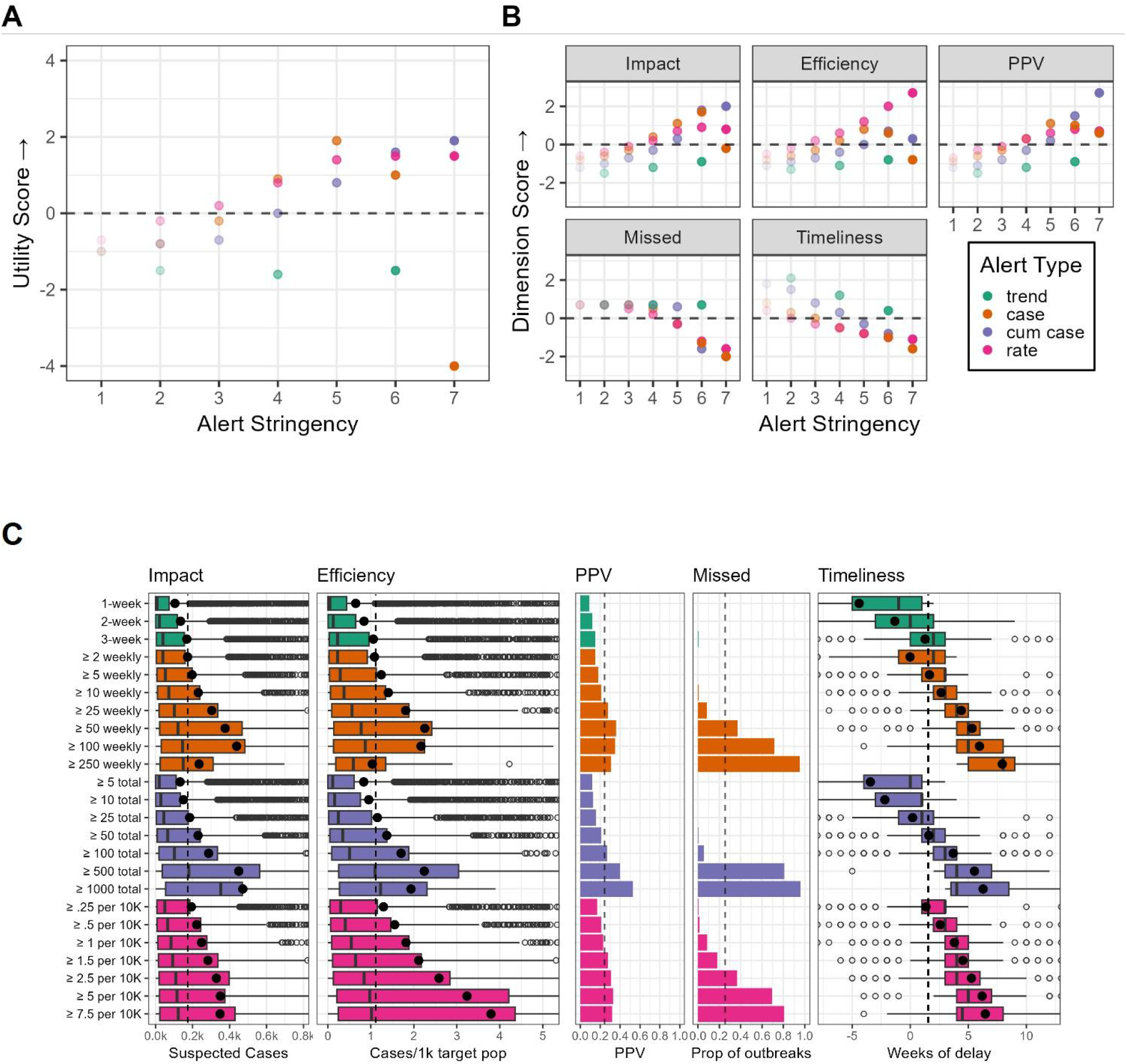
Descriptive distributions of total and dimension utility scores, alert stringency, and dimension metrics by alert definition and type. The scatterplots summarizes the relationship between alert stringency (x-axis and transparency) and A) total utility score (y-axis) or B) dimension score (y-axis and sub-panels) for outbreak-prone subnational locations with a population size between 50,000 and 500,000 across 24 alert definitions (points) grouped by type (color). Higher utility scores indicate higher utility. Alert definition stringency increases from 1 to 7. A horizontal dashed line is placed at the 0 score as each dimension score was centered and standardized. C) The dimension metrics (x-axis) summarize outcomes over a 1-year period following alerts for outbreak-prone locations with a population size between 50,000 and 500,000 across 24 alert definitions (y-axis) grouped by type (color). The impact, efficiency, and timeliness panels are depicted with boxplots, with the black dot representing the mean for each alert definition and the black dashed vertical line indicating the mean of the mean across alert definitions. The PPV and missed dimensions are depicted with bar plots, where bar height indicates proportion and the black dashed vertical line indicates the mean proportion across alert definitions. The color of each bar indicates its alert type as reflected in the color legend in panel B.

Trend-based alerts had lower impact, efficiency, and PPV though they missed few outbreaks compared to weekly case-, cumulative case-, and rate-based alerts (Fig 1C). Case-based and cumulative-case based alerts of moderate stringency balanced between maximizing impact, efficiency, and PPV, and missing few outbreaks, with short delays between alert and outbreak start relative to the average across alert definitions.

### Variations on the utility framework

Where data permitted, we applied a variant of the utility framework that layered laboratory confirmed case thresholds into alert definitions. Increasing confirmed case thresholds were positively associated with the impact, efficiency, and PPV metrics for lower stringency weekly case-based and cumulative case-based alerts (e.g., the mean impact of the joint definition “≥ 2 weekly cases for three consecutive weeks and 10 confirmed cases” was higher than “≥ 50 weekly cases for three consecutive weeks and 0 confirmed cases” (Supp Fig. 11-12). There was no apparent effect on these dimensions for higher stringency alert definitions. By contrast, increasing confirmed case thresholds almost always led to a higher proportion of missed outbreaks and less timely alert triggers.

In another variant of the framework, country stratified utility scores were calculated to assess regional heterogeneity in top-performing alert definitions (Fig 2, Supp Fig. 13-14). In locations with 50,000 to 500,000 people, we saw that “≥ 50 weekly cases for 3 consecutive weeks” was the highest utility alert definition for the greatest number of countries (DR Congo, Ethiopia, Kenya, Tanzania, and Malawi). The definition “≥ 100 cumulative cases over 3 consecutive weeks” was most frequently among the top-3 utility scores, including countries in all four sub-regions of Africa (Chad, Ethiopia, Somalia, South Sudan, Tanzania, Mozambique, Guinea, Niger, Nigeria, Sierra Leone).

**Figure 2.**
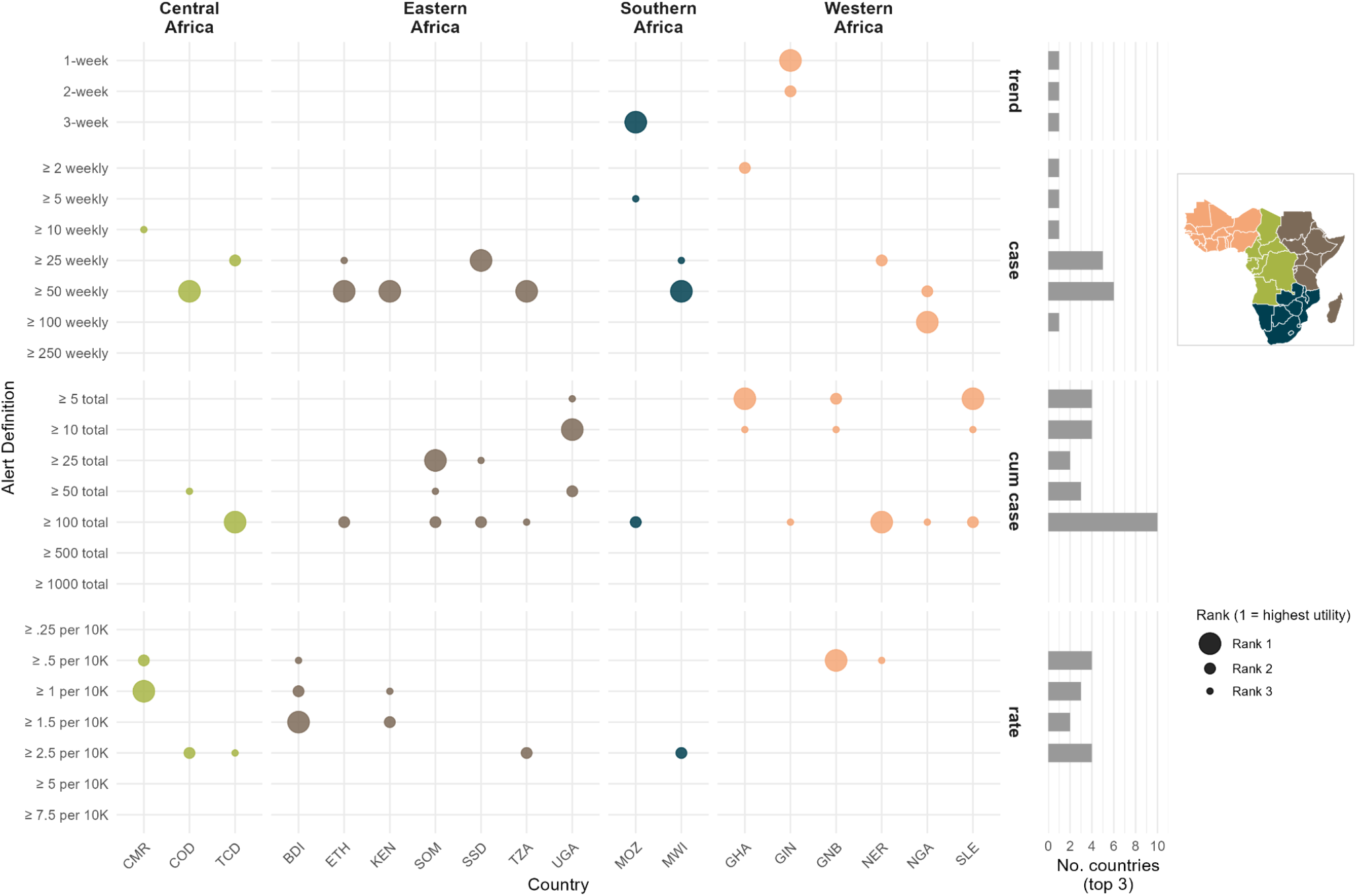
Top three alerts by utility score across countries. Country-specific utility scores were calculated for locations with 50,000 to 500,000 people. Color represents the subregion of Africa, bubble size represents the utility score country rank. The y-axis displays alert definitions grouped into four alert types: trend-based (1-week, 2-week, and 3-week representing exceeding the past 4 week average for x consecutive weeks); case (weekly case thresholds ranging from ≥2 to ≥250 cases per week for three consecutive weeks); cum case (cumulative case thresholds ranging from ≥5 to ≥1,000 total cases for three consecutive weeks); and rate (incidence rate thresholds ranging from ≥0.25 to ≥7.5 cases per 10,000 population for three consecutive weeks). The bar charts to the right of the plot show the number of times each alert definition appeared in a country’s top three alerts definitions based on its country utility score.

### Generalizability of top-performing alert definitions

We sought to identify which alert definitions performed best, that is — had high total utility scores and no low-scoring dimension scores, across pooled and country stratified utility scores — to assess concordance among top-performing definitions across different population contexts. We found that “≥ 50 weekly cases for 3 consecutive weeks” performed well in both pooled and country stratified analyses in populations larger than 50,000 people (Fig 3). In administrative units with 50,000 to 500,000 people, this alert definition had a mean impact of 376 suspected cases (standard deviation: 618), a mean efficiency of 2 (sd: 4.1) cases per 1000 target population, and a mean PPV of 40% (out of 265 alerts), missed a mean of 40% (out of 278) of outbreaks, occurred a mean of 5 weeks (sd: 5) after outbreak start, and had a median 19.8% (95% CrI: −22.9 to −16.9) decrease in the potential impact of interventions for every week of delay (Table 2). In administrative units with over 500,000 people, “≥ 50 weekly cases for 3 consecutive weeks” had a mean impact of 1711 suspected cases (sd: 2650), a mean efficiency of 0.9 (sd: 1.9) cases per 1000 target population, and a mean PPV of 60% (out of 223 alerts), missed a mean of 20% (out of 207) of outbreaks, occurred a mean of 4.2 weeks (sd: 4.6) after outbreak start, and had a median −18.5% (95% CrI: −24.1 to −16.9) change in the potential impact of interventions for every week of delay (Table 2). Cumulative case alert definitions had higher utility for locations with <50,000 people, but the best threshold varied by location type (Fig 3). Trend-based alerts (1-week) were also effective on small populations.

**Figure 3.**
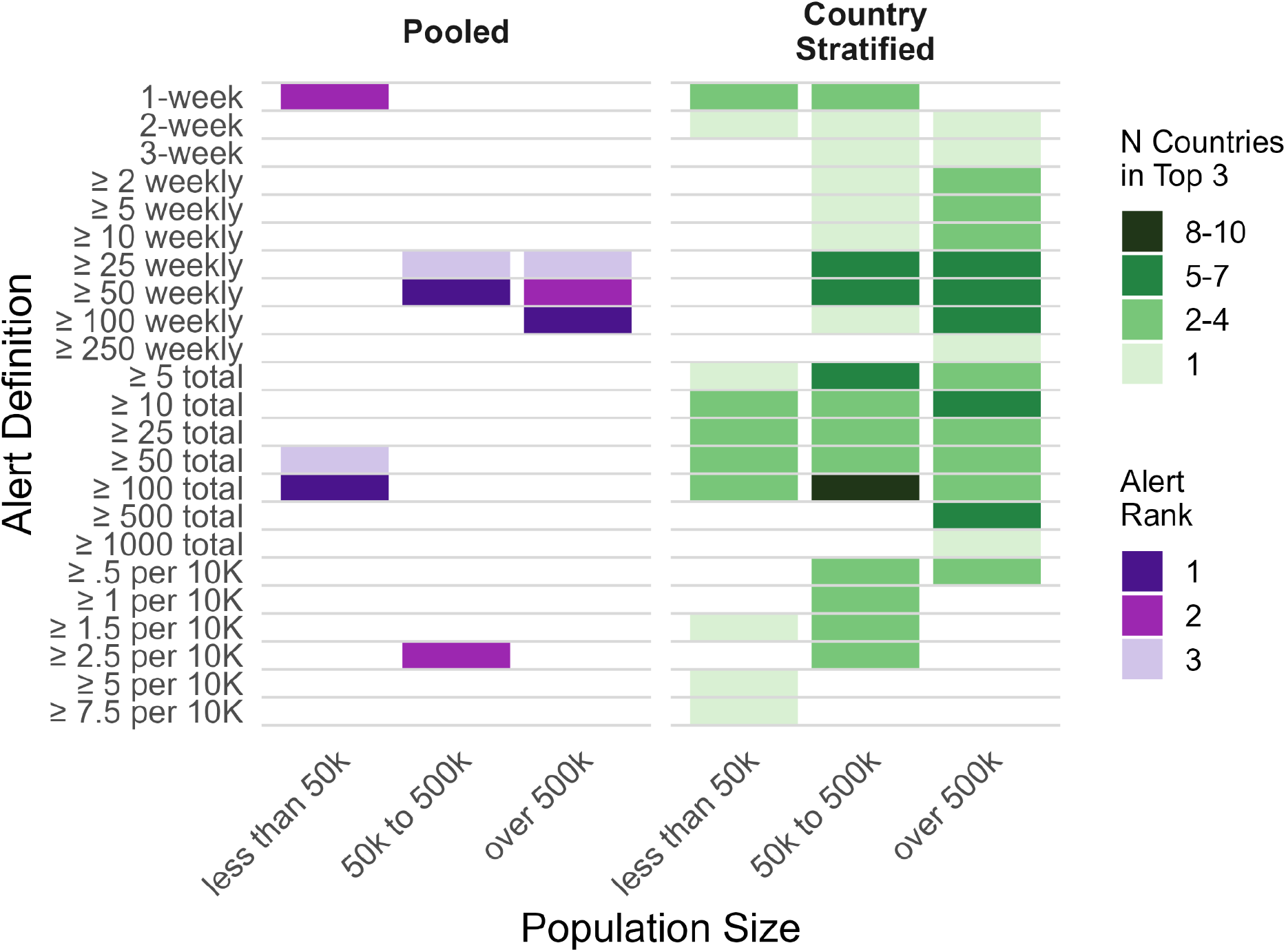
Alert definitions and the transmission context and population group in which they were the top performers. Top performers for the pooled context (left) are the highest utility alerts that have no single dimension score below the cutoff. For the country stratified context (right), color represents the number of countries where the alert definition is in the top-3 for that country, with high values suggesting greater generalizability across countries. Counts are out of the number of countries with a complete utility score in each population group: up to 6 countries in the <50k population group, 18 countries in the 50k–500k group, and 19 countries in the ≥500k; this represents the maximum number of countries that can be ranked in a given population group. A complete utility score requires all five dimensions (impact, efficiency, PPV, missed proportion, and timeliness), and the missed and timeliness dimensions require the country to have experienced at least one linked outbreak in that population band; countries without such outbreaks cannot be ranked. Because alert definitions were not necessarily triggered in every country, individual definitions may not reach these maximum values.

**Table 2.** Top-performing alert definitions for outbreak-prone locations in Africa, evaluated by overall utility score and generalizability across African countries. The table summarizes mean quantitative metrics across five utility dimensions (potential impact, efficiency, PPV, missed, timeliness) and the median estimated percentage change in impact per week of implementation delay for the highest utility alert definitions in outbreak-prone locations. We use the following shorthands for alert types: “trend” for trend-based, “case” for weekly case-based, “cum case” for cumulative case-based, and “rate” for weekly rate-based alerts.

| **Alert Definition** | **Alert Type** | **Potential Impact: cases (SD)** | **Potential Efficiency: cases per 1000 target population (SD)** | **PPV: proportion of alerts (N alerts)** | **Missed Outbreaks: proportion missed by alert (N outbreaks)** | **Timeliness: weeks from outbreak start (SD)** | **Effect of Delay (%): Posterior Median (95 % CrI)** |
| --- | --- | --- | --- | --- | --- | --- | --- |
| **Administrative units with <50,000 people** |  |  |  |  |  |  |  |
| ≥ 100 total | cum case | 274.6 (405.8) | 1031.4 (6928.3) | 0.3 (91) | 0 (43) | 3.3 (4.3) | -19.2 (-24,-14.8) |
| 1-week | trend | 91.4 (260.4) | 234.8 (3257.9) | 0.1 (772) | 0 (43) | -7.6 (15.9) | -10.6 (-13.4,-7.8) |
| ≥ 50 total | cum case | 216.1 (384.7) | 649.5 (5585.1) | 0.2 (149) | 0.2 (149) | 1.6 (2.6) | -17.9 (-22,-14.2) |
| **Administrative units with 50,000 to 500,000 people** |  |  |  |  |  |  |  |
| ≥ 50 weekly | case | 376.2 (618.3) | 2.3 (4.1) | 0.4 (265) | 0.4 (278) | 5.3 (5) | -19.8 (-22.9,-16.9) |
| ≥ 2.5 per 10K | rate | 328.8 (570.8) | 2.6 (4.7) | 0.3 (315) | 0.4 (278) | 5.3 (4.7) | -18.4 (-21.5,-15.4) |
| ≥ 25 weekly | case | 303.8 (550.4) | 1.8 (3.6) | 0.3 (549) | 0.1 (278) | 4.4 (4.5) | -17.8 (-20,-15.7) |
| **Administrative units with ≥500,000 people** |  |  |  |  |  |  |  |
| ≥ 100 weekly | case | 2007.4 (2754.3) | 1.1 (2.1) | 0.7 (157) | 0.4 (207) | 5.8 (5.7) | -21.1 (-25.3,-17.2) |
| ≥ 50 weekly | case | 1711.3 (2650) | 0.9 (1.9) | 0.6 (223) | 0.2 (207) | 4.2 (4.6) | -20.3 (-24.1,-16.9) |
| ≥ 25 weekly | case | 1412.1 (2379.1) | 0.8 (1.8) | 0.6 (317) | 0 (207) | 2.2 (6.6) | -18.5 (-21.6,-15.5) |

### Public access

Users may explore how alert definitions perform across utility dimensions and visualize what happens after an alert with retrospective cholera surveillance data in our public web application accessible at https://diseasedynamics.ch/public/app/cholera_alerts_explorer. One view enables examination of summary statistics of dimension metrics for top-performing alerts across different population and country contexts. A second view visualizes historical time series with select alert definitions so that users may develop intuition about what may happen in the future of ongoing outbreaks.

### Potential change in impact due to implementation delays

The potential impact of an alert depends not only on how rapidly and reliably it detects an outbreak, but also on how quickly action is taken. Thus, for the top-performing alert definition in each population size group, we estimated the effect of implementation delays on the potential impact of an intervention, hypothesizing that long delays would be associated with a smaller impact in the 1-year post-alert evaluation period. For every week of implementation delay, the top-performing definitions corresponded to a 19 to 21% decrease in potential impact (<50,000 people: −19%, 95% CrI: −24 to −15; 50,000 to 500,000 people: −20%, 95% CrI: −23 to −17; ≥500,000 people: −21%; 95% CrI: −24 to −17) (Table 2, Supp Tab. 8-10, Supp Fig. 15). The Democratic Republic of the Congo was notably less sensitive to implementation delays than in other countries, particularly for locations with 50,000 to 500,000 people (−11%; 95% CrI: −13 to −9) (Fig 4, Supp Fig. 16). Other country-specific delay impact estimates had overlapping 95% CrIs with the global delay impact estimate and there was large uncertainty in the posterior distributions.

**Figure 4.**
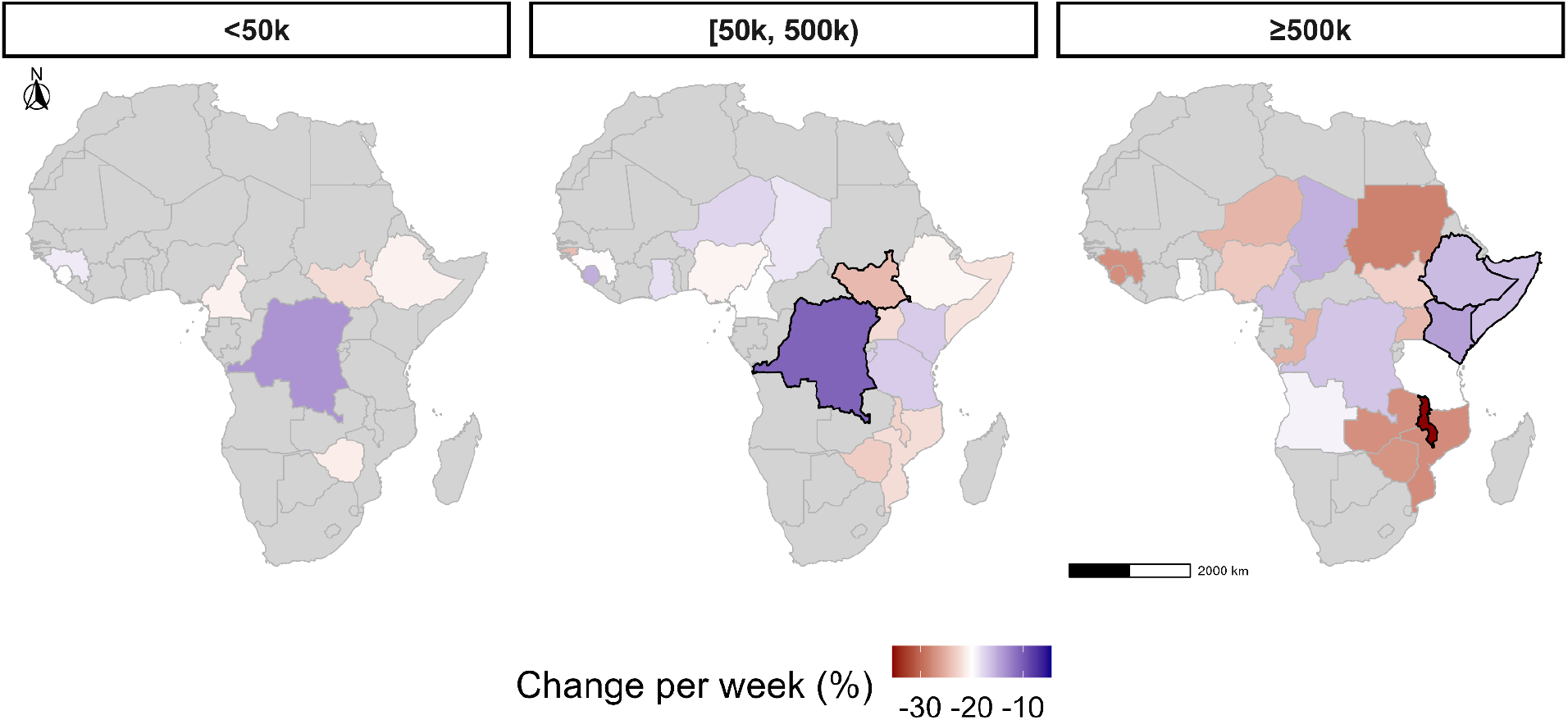
The estimated median percent change per week of implementation delay for the highest utility alert across countries and population groups. Countries are outlined with a dark black border only if the 95% CrI of the posterior for the country-specific deviation does not overlap with 0% (indicating no significant deviation from the global estimated slope). Countries are filled grey if no alerts were triggered. The color scale is centered on the global median estimate (white color) for the population group with 50,000 to 500,000 people.

## Discussion

We developed a decision analytic framework that can help decision makers determine the magnitude of their response after initial signals of cholera activity. The multidimensional nature of the framework enables consideration of tradeoffs between different utility dimensions and manage expectations about whether early case-based signals will expand into larger cholera outbreaks and how large those outbreaks will be. In applying our framework to outbreak-prone settings in Africa, we found that simple, heuristically-defined thresholds based on suspected cholera case counts, while imperfect, can serve as actionable triggers for large-scale interventions like reactive vaccination campaigns across multiple epidemiologic settings. Our simple web-based application can provide real-time support to decision makers in setting expectations about future cholera activity after an alert is triggered.

Two alert definitions performed reasonably well across utility dimensions and should be considered as triggers for large-scale interventions in many contexts, particularly when only suspected case surveillance is available; “≥ 50 weekly cases for three consecutive weeks” performed best in populations over 50,000, while “≥ 100 total cases over three consecutive weeks” performed well in all population sizes in many countries. Overall, weekly case-based alerts performed better in larger populations, while cumulative case-based alerts were better adapted to smaller populations and contexts with reporting gaps. When we assessed the robustness of the potential impact of these top-performing alert definitions to intervention implementation delays, we found that every week of delay represented a roughly 20% decrease in the potential impact of an intervention. There were, however, some meaningful deviations from this continent-wide average (e.g., potential impact in DR Congo was less sensitive to delay), suggesting that the local epidemiological context shapes not only which alert definitions perform best, but also the relative importance of a timely and decisive response.

Our analysis provides a useful baseline expectation, but we acknowledge that a limited set of utility framework configurations (sum of centered and standardized dimension score vs. rank-sum-based score; dimension score threshold for exclusion from top-performing alert definition list), evaluation approaches (pooled vs. country stratified), and alert definitions (trend-based, weekly and cumulative case-based, weekly rate-based, with and without confirmed case thresholds) were explored. Rather than promoting our specific implementation as the optimal approach, we propose that others borrow from our conceptual framing – the idea of combining utility dimensions like impact, positive predictive value, and timeliness into a multidimensional score and evaluating potential signals from an early, decision-making window – to identify alert definitions appropriate to their context and priorities. For example, countries may customize the framework to use their own surveillance data as a historical baseline and weigh dimensions according to their national priorities. Indeed, in situations where daily case surveillance, laboratory confirmation, or other cholera risk factor data are available, alert definitions should be reconsidered to leverage this information. Such improvements may bring cholera outbreak response closer to the “7-1-7” global health security target: seven days to detect emerging outbreaks, one day to notify public health authorities, and seven days to implement early warning response [22,23].

While we used the largest available cholera surveillance time series dataset known to us to develop our approach, a more generalizable evaluation of this and other alert frameworks is warranted. Future work should seek to evaluate alert definitions with globally-representative cholera surveillance data, and with a model that accounts for locations that were vaccinated before or during an outbreak to account for extrinsic population immunity, should such data become available. A greater volume of data should also be sought for cholera-endemic locations, so that a retrospective evaluation of a more stringent set of alert definitions could be performed for higher burden areas in the same utility framework. We also recommend that different types of alerts be considered for future analyses – 40% of outbreaks over 300 cases were still missed when applying our top-performing definitions, and it is possible that different variations of case-based signals (e.g., exceeding a weekly case threshold or a cumulative case threshold) could better capture the breadth of predictive outbreak signals.

As support for global health faces broad financial cuts, evidence-based anticipatory action, a risk management approach which relies on pre-agreed-upon triggers for humanitarian and emergency response, is increasingly important for public health decision making under uncertainty [24,25]. Our decision analytic framework and publicly-accessible web application fill an operational gap in cholera emergency response and our top-performing alert definitions could be piloted as decision triggers for submitting or approving requests to the OCV stockpile or other emergency funding and resources. Because our approach requires no new data streams or advanced algorithms at baseline, relying only on suspected case surveillance, it can be integrated directly into existing cholera surveillance frameworks to make decisions about rapid resource mobilization in affected locations quickly and efficiently.

## Supporting information

Supplemental Results

## Data Availability

All data produced in the present study upon reasonable request to the authors.

https://doi.org/10.12688/gatesopenres.16378.1

https://osf.io/2ncf7

## Competing interests

ASA and ECL participate in the Global Task Force on Cholera Control Surveillance and Oral Cholera Vaccine Working groups, which provide technical expertise on cholera surveillance and oral cholera vaccine use. ASA is a member of the Gavi Independent Review Committee.

## Funding

This work was carried out as part of the Vaccine Impact Modelling Consortium (www.vaccineimpact.org), but the views expressed are those of the authors and not necessarily those of the Consortium or its funders. This work was supported, in whole or in part, by the Gates Foundation, via the Vaccine Impact Modelling Consortium [Grant Number INV-034281], previously (OPP1157270 / INV-009125) and Gavi, the Vaccine Alliance. The conclusions and opinions expressed in this work are those of the author(s) alone and shall not be attributed to the Foundation. Under the grant conditions of the Foundation, a Creative Commons Attribution 4.0 License has already been assigned to the Author Accepted Manuscript version that might arise from this submission.

## Notes

### Author Declarations

According to the Institutional Review Board (IRB) at the Johns Hopkins Bloomberg School of Public Health (BSPH), the surveillance and outbreak datasets constructed from the Cholera Taxonomy database were exempt (BSPH IRB No. 27682).

