## Supplemental Methods for "A decision analytic framework for triggering cholera outbreak response based on early-case surveillance"

### Supplementary Methods

#### Data processing figures

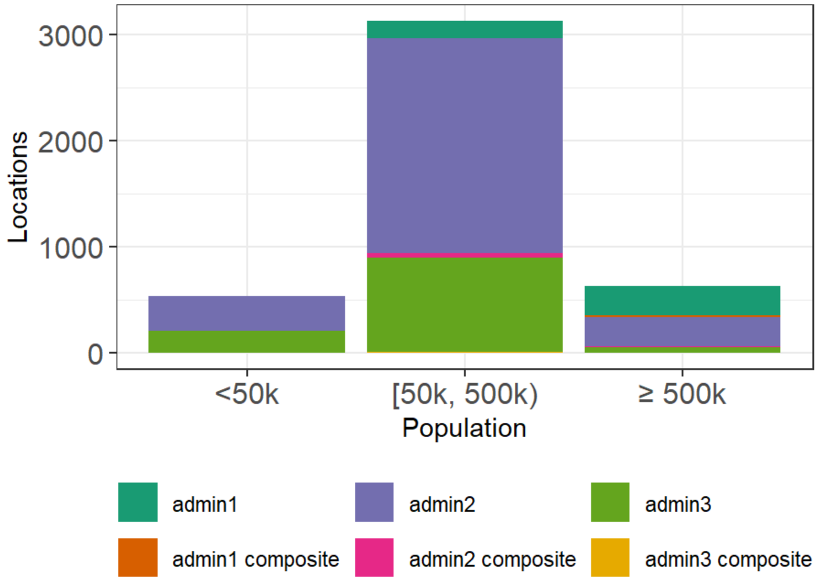


**Supp. Fig. 1: Population distribution of location-population units by administrative unit scale for all outbreak-prone locations represented in the weekly consecutive time series.** "Composite" locations are those representing the union of multiple administrative units (e.g., admin1 composite represents the combined data of multiple administrative level 1 units all together).

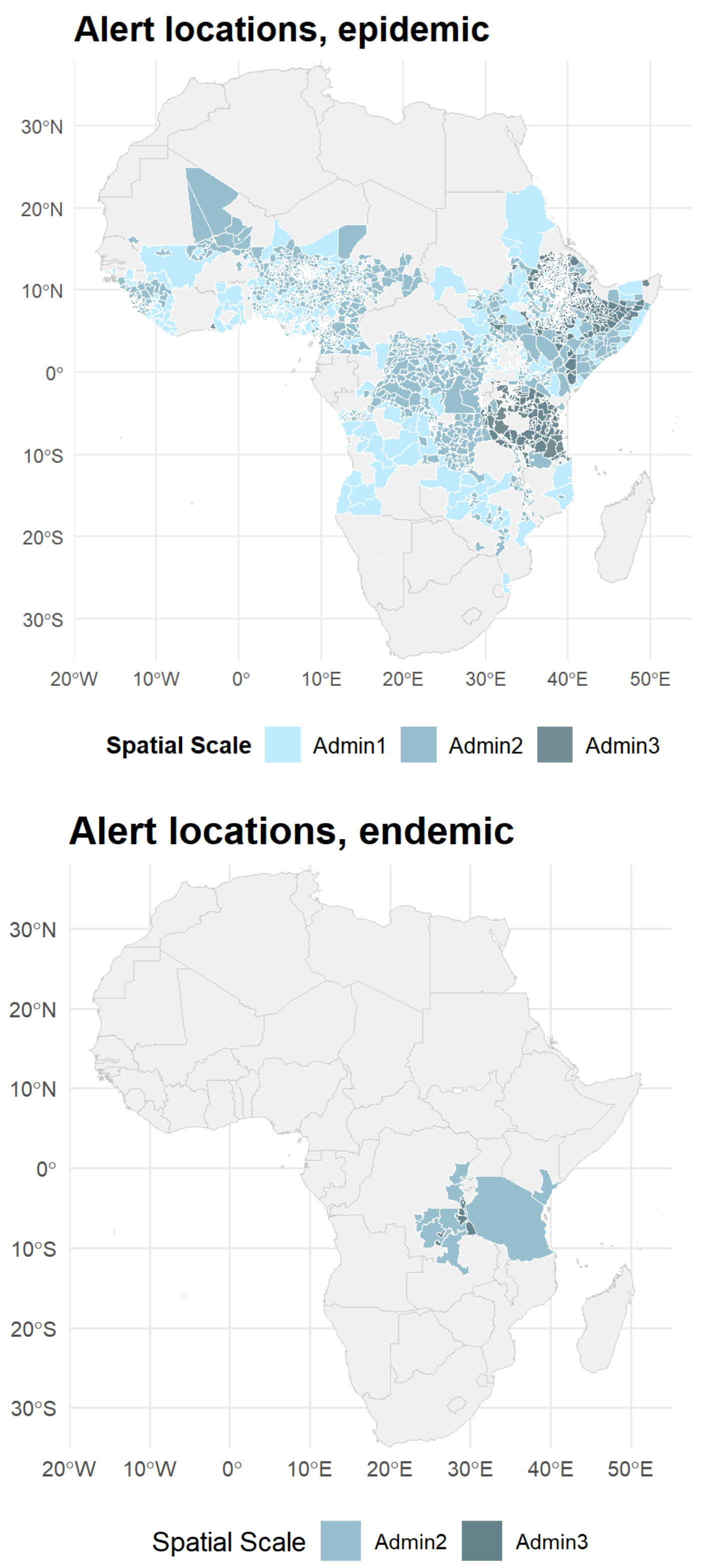


**Supp. Fig. 2: Maps with outbreak prone alert locations (upper panel, n = 2592) and endemic alert locations (lower panel, n = 46) in Africa, colored by administrative level.** Alerts in endemic locations were excluded from the analysis.

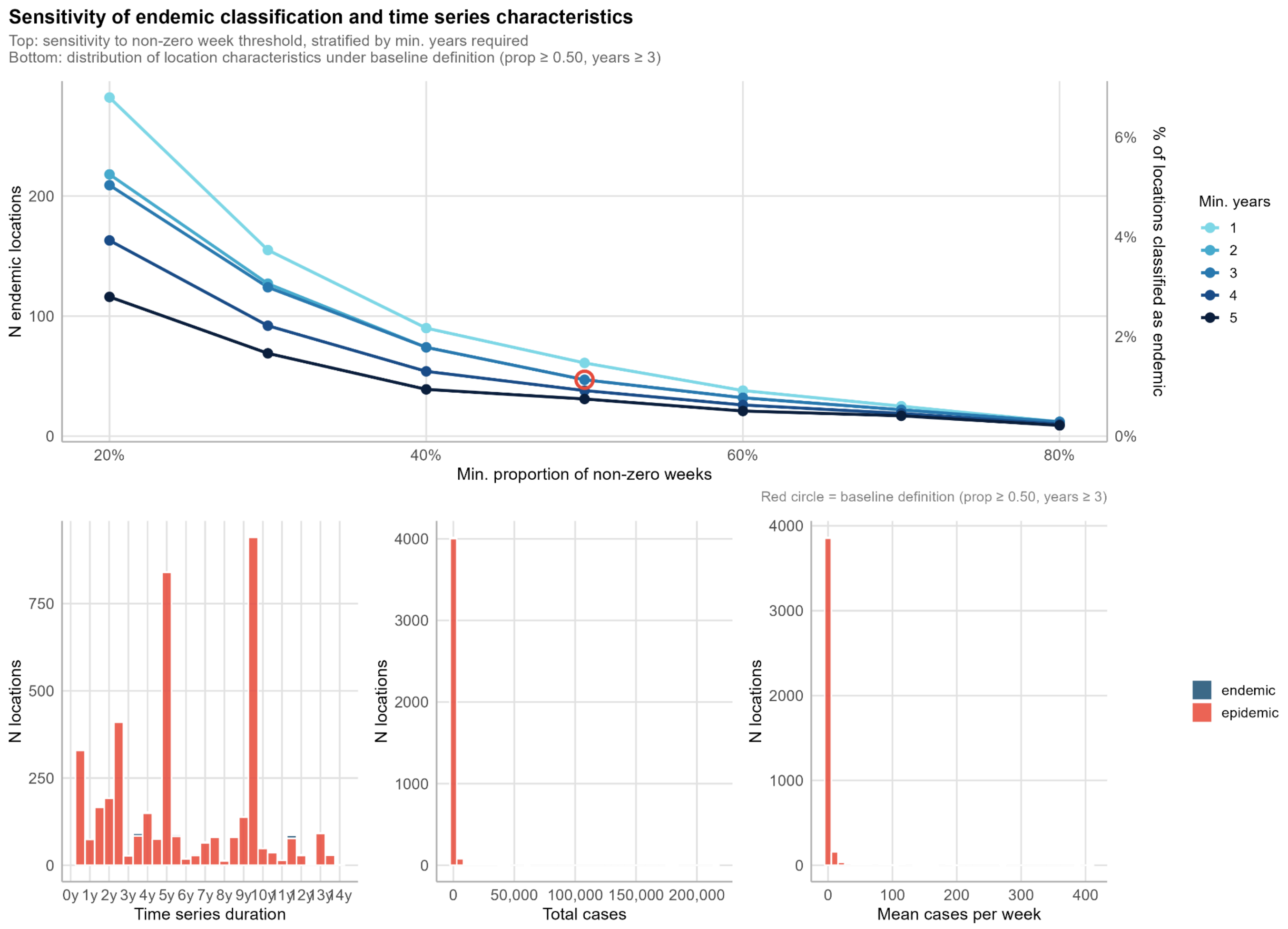


**Supp. Fig. 3: Comparison of multiple potential definitions of endemic locations and their respective characteristics.** Top: number (left axis) and proportion (right axis) of locations classified as endemic across combinations of the minimum proportion of non-zero case weeks (x-axis) and minimum years of data required (line colour). The red circle marks the baseline definition (≥ 50% non-zero weeks, ≥ 3 years of data). Bottom: distribution of time series duration (left), total reported cases (centre), and mean weekly cases (right) across all locations under the baseline endemic definition. Blue = endemic locations; red = epidemic locations.

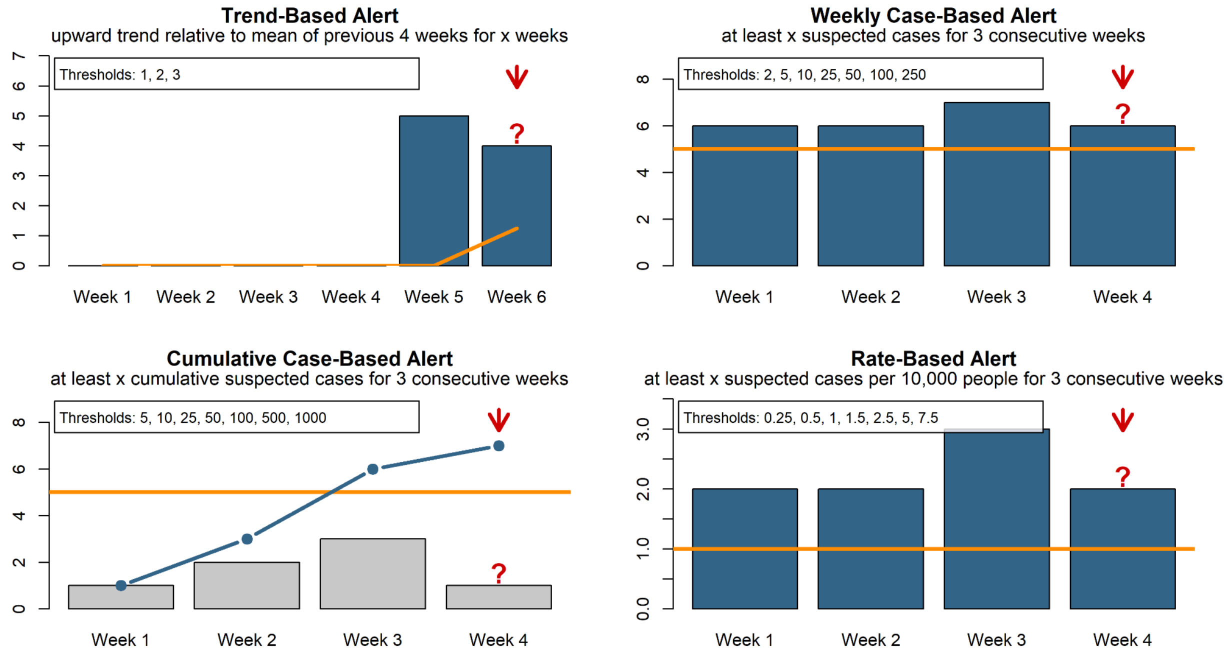


**Supp. Fig. 4: Conceptual representation of the four alert types.** The panels depict trend-based (trend), weekly case-based (case), cumulative case-cased (cum case), and rate-based alerts (rate). Alerts are triggered (red arrow) the week after the threshold condition (described in panel subtitles) has been met. Weekly bar color is grey when the threshold has not been exceeded and blue when it has been exceeded. For cumulative case-based alerts, the blue line indicates the number of cumulative cases over the four week period. Orange lines indicate an example threshold that must be exceeded to trigger the alert. The threshold values for each alert type are embedded in each figure.

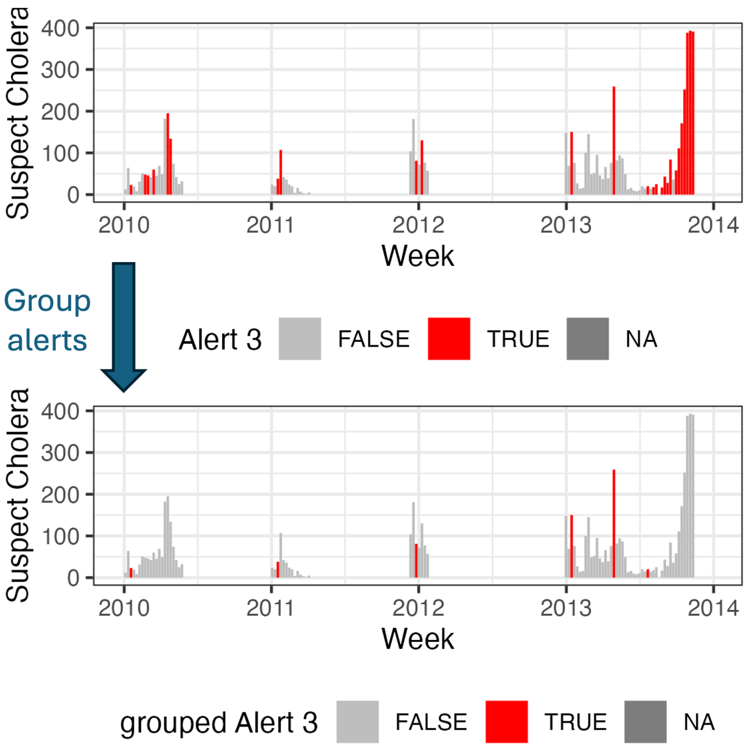


**Supp. Fig. 5: Conceptual diagram on how temporally-clustered alerts are grouped together until an 8-week reset period of no alerts is achieved.** Since an alert would lead to a period of heightened public health awareness, temporally-proximate alerts in the same location were grouped together until 8 consecutive weeks without an alert were observed and the period to search for new alerts was "reset." The term "alert" refers to the date of the first alert in a series of temporally-grouped alerts.

### Alert definitions

Different alert types were defined to capture heterogeneity in transmission and reporting dynamics. Trend-based alerts aim to capture transmission patterns that arise from exceeding a baseline or seasonal threshold and use moving averages, widely used for influenza outbreak detection (e.g. ref). Weekly case-based alerts required threshold exceedance for three consecutive weeks to identify sustained transmission, while cumulative case-based alerts identified gradual increases by aggregating cases over three weeks. These approaches build on GTFCC suspected-case threshold guidance while incorporating temporal persistence and cumulative burden, using simple rounded thresholds to support operational decision-making and alignment with current guidance. Cumulative case-based alerts definitions are less stringent than weekly case-based alerts because they are less sensitive to temporal gaps in reporting. Finally, rate-based alerts mirror case-based alerts but use incidence rates to ensure comparability across populations of different sizes.

Temporally-clustered alerts were grouped together to avoid triggering multiple alerts in the same apparent outbreak (Supp Fig. 5). The alert date for an alert was recorded as the date of the first alert in a series of temporally-clustered alerts. Subsequent alerts triggered in the same definition were grouped under the initial alert until 8 consecutive weeks without an alert were observed, after which the search for a new alert could begin again. The motivation for grouping together temporally-clustered alerts is to avoid including highly temporally autocorrelated results (e.g., metrics representing only a 1-week frameshift after a given alert) in our dimension metric calculations.

#### Alert definitions enhanced with case confirmation

To explore the effect of combining alert signals (based on suspected cases) with case confirmation on alert utility, we repeat the alert utility analysis for subnational outbreak-prone locations with populations from 50,000 to 500,000 people, keeping only alerts that had a certain number of confirmed cases in the alert period (the week of the alert and the three weeks preceding it). The analysis was applied only to weekly case-based, cumulative case-based, and rate-based alerts and definitions with stringency level 5 or lower as a proof-of-concept about the effects of adding case confirmation thresholds to more sensitive alert definitions which may have low utility due to a high “false-alert” rate (N.B. a case confirmation threshold of 0 corresponds to suspected case alerts reported in the primary text). We hypothesize that adding case confirmation thresholds will reduce the false-alert rate, thus increasing utility for high sensitivity / low stringency definitions. We repeat the analysis for different thresholds of confirmed cases. As an additional sensitivity analysis, we compare dimension metrics for low stringency alert definitions of the case-based type at different case confirmation thresholds to the top-performing case-based alert definition at a 0 case confirmation threshold in locations with 50,000 to 500,000 people (Supp Fig 9).

To recalculate the missed and timeliness dimensions for this secondary analysis, we linked the filtered alerts to the subset of outbreaks linked to confirmed case alerts at the given confirmed case threshold threshold. There was limited case confirmation data and most alerts with at least 1 confirmed case were clustered in Ethiopia (Supp Fig 5).

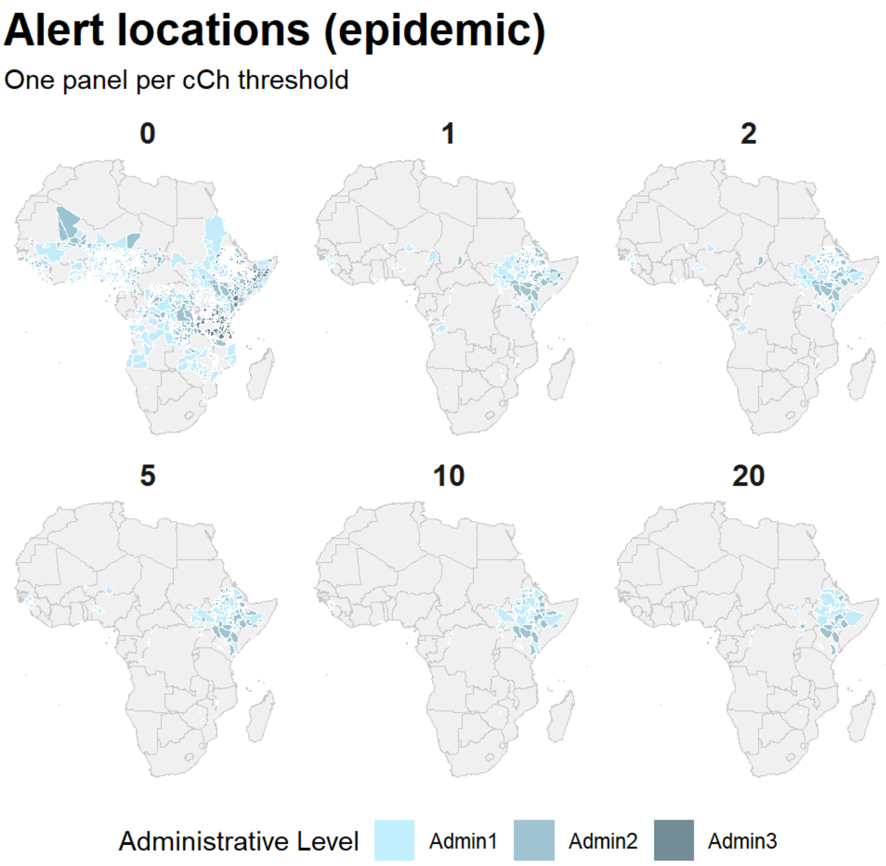


**Supp. Fig. 6: Alert locations with available confirmed cases (cCh) during the alert period according to different thresholds.**

#### Efficiency dimension metric

Efficiency was defined as the ratio of reported suspected cases in the 1-year post-alert evaluation period to the target population of a potential OCV campaign (all individuals aged 1 year and above). Target populations were calculated for each location as the product of the UN-adjusted WorldPop estimates (see main text) and annual country-level population estimates of the proportion of the population aged 1-year and above according to the United Nations World Population Prospects.

#### Missed and timeliness dimension metrics

Categorizing which outbreaks are missed and the timeliness of an alert relative to an outbreak start requires us to compare our alerts to a separate outbreak definition. To this end, we leveraged the cholera outbreak dataset extracted in Zheng et al's (2026), which used the same underlying surveillance database but defines the outbreak start and end with an approach that can only be applied retrospectively. In brief, outbreaks began when the weekly cholera incidence rate exceeded a location-specific threshold for at least two consecutive weeks and ended after two weeks below the threshold when followed by a four-week washout.

For all alert definitions, we attempted to link each outbreak with a unique alert definition instance. Following an algorithmic approach, we identified the earliest alert (period of heightened alert activity) whose temporal interval overlapped or shortly preceded the outbreak period (Supp Fig 6). Overlapping alerts were defined as those triggered earlier or on the same date as the end of the outbreak and ended later or on the same date as the start of the outbreak. We also identified the alert triggered shortly before each outbreak started (the latest alert whose period ended before the start of the outbreak) and the alert triggered shortly after each outbreak ended (the earliest alert whose period started after the outbreak ended). In a subsequent filtering step, we selected only the earliest overlapping alert for each outbreak and in the rare cases where there was no overlapping alert, we manually inspected our outbreak/alert time series data and selected the alert triggered shortly before each outbreak. All linkages were validated with manual inspection.

When no alert overlapped an outbreak, the outbreak was considered to be “missed” by the given alert definition. The timeliness metric was calculated as the difference between the alert period start and the outbreak start dates.

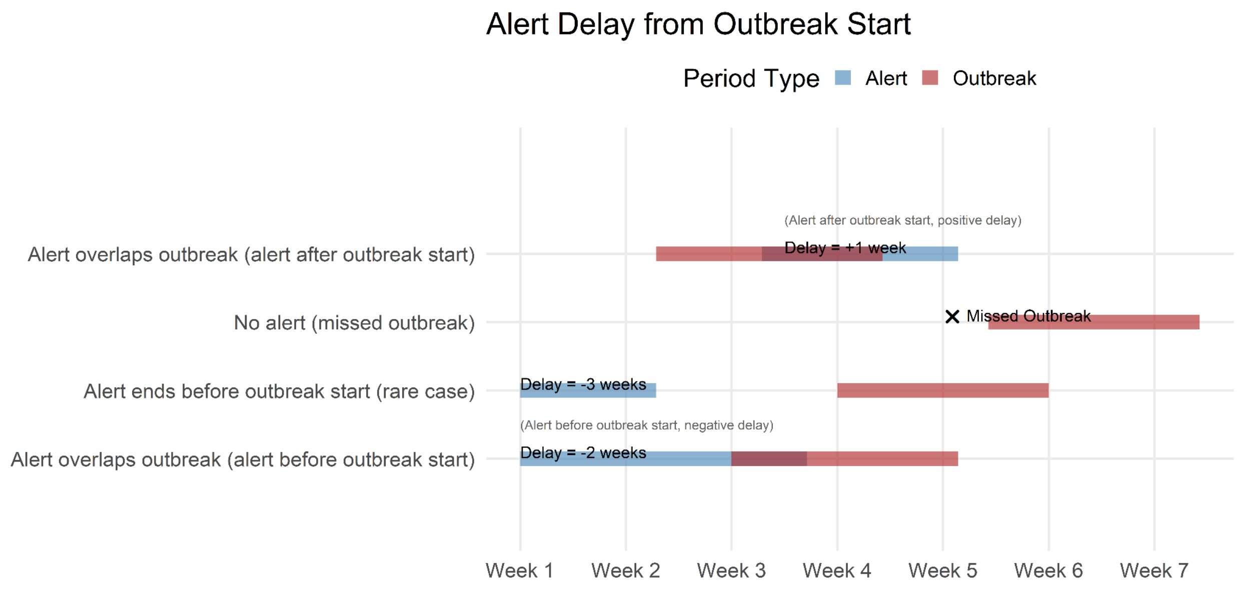


**Supp. Fig. 7: The procedure used to calculate the metric representing the timeliness of alerts relative to outbreak periods.** Positive and negative delay values are possible when alerts are triggered after the outbreak start (top row) and when alerts are triggered before the outbreak start respectively (bottom two rows).

#### Choice in dimension score thresholds

Top-performing alert definitions were identified as those with the highest utility scores and no dimension score below a threshold value of -1 (i.e., no dimension had a very poor performance). We explored several dimension score thresholds and were motivated to select the threshold of -1 because it was the least restrictive threshold that enabled reasonably good performance across all dimension metrics on their original scales (Supp Tabs 5-6).

**Supp Tab 1: Top 4 alerts by utility score per population group, where no utility dimension is below the cutoff value equal to -0.8.**

| Alert Definition | Alert Type | Impact: cases (SD) | Efficiency: cases per 1000 target population (SD) | PPV: proportion of alerts (N alerts) | Missed: proportion of outbreaks (N outbreaks) | Delay: weeks from outbreak start (SD) | Utility Score |
|---|---|---|---|---|---|---|---|
| **Administrative units with <50,000 people** | | | | | | | |
| **≥ 100 total** | **cum case** | **274.6 (405.8)** | **1031.4 (6928.3)** | **0.3 (91)** | **0 (43)** | **3.3 (4.3)** | **2.4** |
| ≥ 50 total | cum case | 216.1 (384.7) | 649.5 (5585.1) | 0.2 (149) | 0 (43) | 1.6 (2.6) | 1.1 |
| ≥ 10 weekly | case | 204.4 (352.6) | 652.1 (5485.5) | 0.2 (139) | 0 (43) | 2.7 (2.3) | 0.6 |
| ≥ 25 total | cum case | 182.4 (392.7) | 446.6 (4693.9) | 0.1 (233) | 0 (43) | 0.3 (2.3) | 0.2 |
| **Administrative units with 50,000 to 500,000 people** | | | | | | | |
| **≥ 50 weekly** | **case** | **376.2 (618.3)** | **2.3 (4.1)** | **0.4 (265)** | **0.4 (278)** | **5.3 (5)** | **1.9** |
| ≥ 2.5 per 10K | rate | 328.8 (570.8) | 2.6 (4.7) | 0.3 (315) | 0.4 (278) | 5.3 (4.7) | 1.4 |
| ≥ 25 weekly | case | 303.8 (550.4) | 1.8 (3.6) | 0.3 (549) | 0.1 (278) | 4.4 (4.5) | 0.9 |
| ≥ 100 total | cum case | 286.9 (527.9) | 1.7 (3.5) | 0.3 (723) | 0.1 (278) | 3.7 (5.2) | 0.8 |
| **Administrative units with ≥500,000 people** | | | | | | | |
| **≥ 100 weekly** | **case** | **2007.4 (2754.3)** | **1.1 (2.1)** | **0.7 (157)** | **0.4 (207)** | **5.8 (5.7)** | **1.0** |
| ≥ 50 weekly | case | 1711.3 (2650) | 0.9 (1.9) | 0.6 (223) | 0.2 (207) | 4.2 (4.6) | 0.8 |
| ≥ 25 weekly | case | 1412.1 (2379.1) | 0.8 (1.8) | 0.6 (317) | 0 (207) | 2.2 (6.6) | 0.6 |
| ≥ 100 total | cum case | 1342.5 (2406) | 0.8 (1.8) | 0.5 (389) | 0 (207) | 0.7 (7.3) | 0.5 |

**Supp Tab 2: Top 4 alerts by utility score per population group, where no utility dimension is below the cutoff value equal to -0.6.**

| Alert Definition | Alert Type | Impact: cases (SD) | Efficiency: cases per 1000 target population (SD) | PPV: proportion of alerts (N alerts) | Missed: proportion of outbreaks (N outbreaks) | Delay: weeks from outbreak start (SD) | Utility Score |
|---|---|---|---|---|---|---|---|
| **Administrative units with <50,000 people** | | | | | | | |
| **≥ 50 total** | **cum case** | **216.1 (384.7)** | **649.5 (5585.1)** | **0.2 (149)** | **0 (43)** | **1.6 (2.6)** | **1.1** |
| ≥ 10 weekly | case | 204.4 (352.6) | 652.1 (5485.5) | 0.2 (139) | 0 (43) | 2.7 (2.3) | 0.6 |
| ≥ 25 total | cum case | 182.4 (392.7) | 446.6 (4693.9) | 0.1 (233) | 0 (43) | 0.3 (2.3) | 0.2 |
| ≥ 5 per 10K | rate | 173.6 (316.9) | 574.7 (5038) | 0.2 (165) | 0 (43) | 2.7 (2.9) | -0.3 |
| **Administrative units with 50,000 to 500,000 people** | | | | | | | |
| **≥ 25 weekly** | **case** | **303.8 (550.4)** | **1.8 (3.6)** | **0.3 (549)** | **0.1 (278)** | **4.4 (4.5)** | **0.9** |
| ≥ 100 total | cum case | 286.9 (527.9) | 1.7 (3.5) | 0.3 (723) | 0.1 (278) | 3.7 (5.2) | 0.8 |
| ≥ 1.5 per 10K | rate | 282.9 (509.3) | 2.1 (4) | 0.3 (500) | 0.2 (278) | 4.5 (4.4) | 0.8 |
| ≥ 1 per 10K | rate | 248.4 (458.1) | 1.8 (3.5) | 0.2 (721) | 0.1 (278) | 3.8 (4) | 0.2 |
| **Administrative units with ≥500,000 people** | | | | | | | |
| **≥ 50 weekly** | **case** | **1711.3 (2650)** | **0.9 (1.9)** | **0.6 (223)** | **0.2 (207)** | **4.2 (4.6)** | **0.8** |
| ≥ 25 weekly | case | 1412.1 (2379.1) | 0.8 (1.8) | 0.6 (317) | 0 (207) | 2.2 (6.6) | 0.6 |
| ≥ 100 total | cum case | 1342.5 (2406) | 0.8 (1.8) | 0.5 (389) | 0 (207) | 0.7 (7.3) | 0.5 |
| ≥ 50 total | cum case | 1057.5 (2171.3) | 0.7 (1.7) | 0.4 (508) | 0 (207) | -2.3 (9.6) | -0.1 |

#### Alternative utility score calculation (rank sum score)

We performed a sensitivity analysis of the utility score calculation by comparing two methods of defining the utility score. The first method is described in the main text. The alternative is a rank sum score. For every dimension within each population group, metrics where higher values indicated better performance (Impact, Efficiency, PPV) were ranked in ascending order, and metrics where lower values indicated better performance (Missed, Delay) were ranked in descending order. We then summed the ranked dimension scores to obtain the rank sum utility score. The association between the two utility score calculations is linear with a positive slope (Supp Fig 10), indicating that both methods yield similar relative rankings in utility, despite being expressed in different scales. The association between the dimension scores between the two approaches is linear and positive (Supp Fig 11). The only population group where there is a slightly non-linear trend between the two calculations is the population group under 50,000 people.

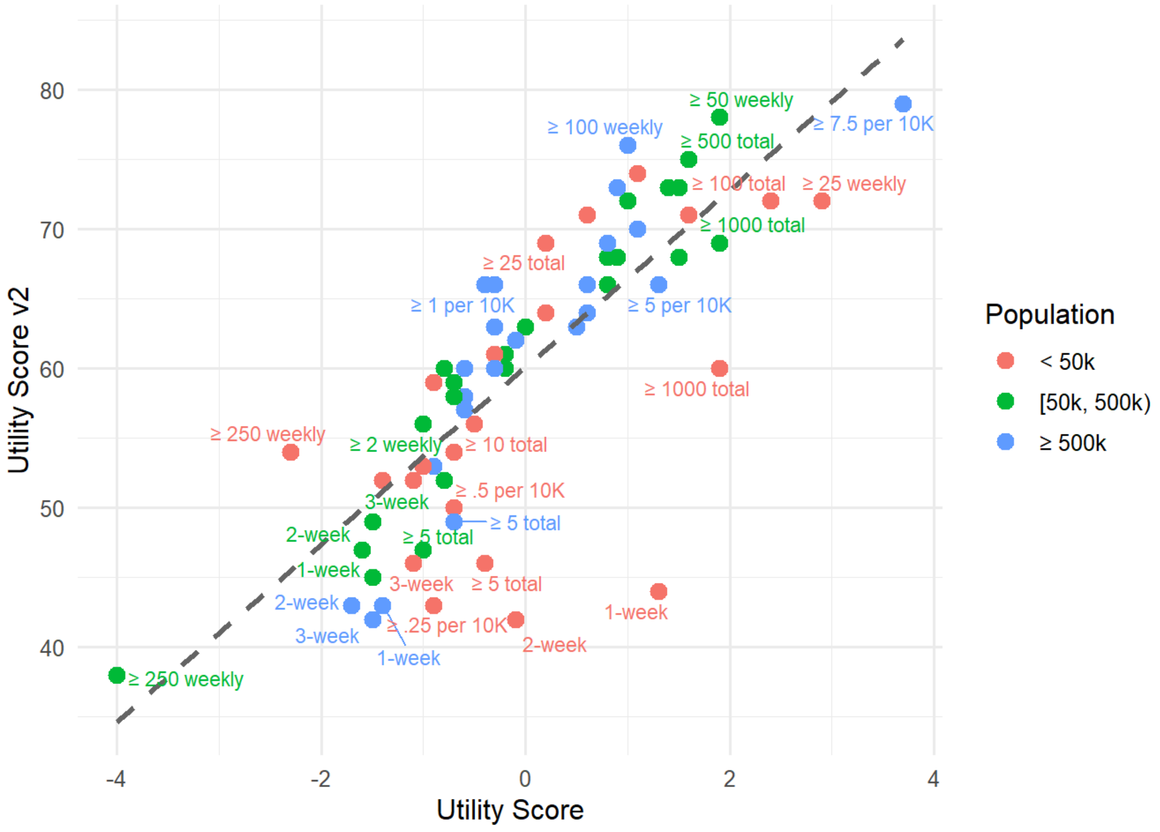


**Supp Fig 8. Scatterplot of the utility score on the x axis and the alternate rank sum utility score (v2) on the y axis.** Color represents the population group.

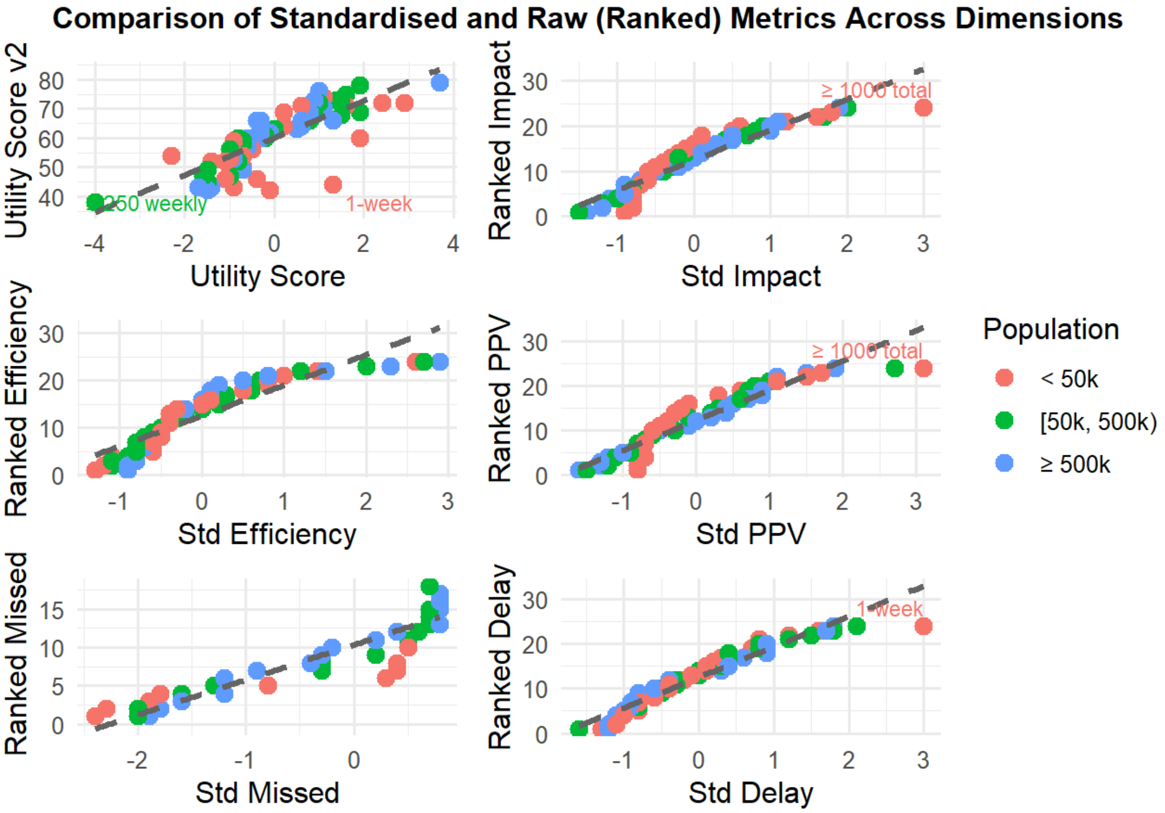


**Supp Fig 9. Scatterplot of the utility score and the standardized dimension scores on the x axis and the alternate rank sum utility score (v2) and the ranked dimension scores and on the y axis.** Color represents the population group.

**Supp Tab 3: Best alert definitions per population group by utility score v2 (rank-sum).** The alert definitions highlighted in green are the selected best alert definitions per population group using the original utility score and standardised dimension scores.

| Alert Definition | Alert Type | Impact: cases | Efficiency: per 1000 pop | PPV | Missed Proportion | Delay (weeks) | Utility Score (V2) | Utility Score (Original) |
|---|---|---|---|---|---|---|---|---|
| **Administrative units with <50,000 people** | | | | | | | | |
| ≥ 50 total | cum case | 216.0678 | 649.501487 | 0.19 | 0.0000000 | 1.627907 | 74 | 1.1 |
| ≥ 25 weekly | case | 286.2338 | 1223.902741 | 0.31 | 0.0232558 | 4.833333 | 72 | 2.9 |
| ≥ 100 total | cum case | 274.6421 | 1031.382599 | 0.26 | 0.0465116 | 3.341463 | 72 | 2.4 |
| ≥ 50 weekly | case | 361.4847 | 855.724599 | 0.37 | 0.4418605 | 4.125000 | 71 | 1.6 |
| **Administrative units with 50,000 to 500,000 people** | | | | | | | | |
| ≥ 50 weekly | case | 376.2355 | 2.255394 | 0.36 | 0.3705036 | 5.325714 | 78 | 1.9 |
| ≥ 500 total | cum case | 449.7266 | 2.243498 | 0.40 | 0.8093525 | 5.528302 | 75 | 1.6 |
| ≥ 5 per 10K | rate | 350.0699 | 3.237352 | 0.33 | 0.6942446 | 6.188235 | 73 | 1.5 |
| ≥ 2.5 per 10K | rate | 328.7691 | 2.585916 | 0.31 | 0.3669065 | 5.272727 | 73 | 1.4 |
| **Administrative units with ≥500,000 people** | | | | | | | | |
| ≥ 7.5 per 10K | rate | 2351.4444 | 3.506429 | 0.89 | 0.9565217 | 6.111111 | 79 | 3.7 |
| ≥ 100 weekly | case | 2007.3553 | 1.128650 | 0.73 | 0.3768116 | 5.767442 | 76 | 1.0 |
| ≥ 1000 total | cum case | 2444.1656 | 1.255141 | 0.74 | 0.7149758 | 7.203390 | 73 | 0.9 |
| ≥ 250 weekly | case | 2484.9370 | 1.189461 | 0.81 | 0.7149758 | 7.966102 | 70 | 1.1 |

### Delay model equations

The main model used to estimate the effect of additional weeks of implementation delay on the potential impact of intervention takes the following form:

$$r_i = log(\frac{y_{d_i}}{y_{delay\ at\ week\ 0}})$$

$$r_i \sim N(\mu_i, \sigma), \qquad \sigma \sim HalfNormal(1)$$

$$\mu_i = \beta_{alert[i]} d_i$$

$$\beta_{alert[i]} = \alpha_{global} + \alpha_{country[i]} + \alpha_{alert[i]}$$

$$\alpha_{global} \sim N(0, \sigma_{global}), \qquad \sigma_{global} \sim HalfNormal(1)$$

$$\alpha_{country} \sim N(0, \sigma_{country}), \qquad \sigma_{country} \sim HalfNormal(0.5)$$

$$\alpha_{alert} \sim N(0, \sigma_{alert}), \qquad \sigma_{alert} \sim HalfNormal(0.25)$$

where $r_i$ is the log-ratio, is measurement variability for the log ratio, $\beta_{alert[i]}$ is the alert-level effect of delay $d_i$ for observation $i$ ,so that the effect of delay is given by a single alert-level slope for multiple observations $i$ at the alert level, each corresponding to a different delay value $d_i$. The alert level effect of delay is made up of the global effect of delay $\alpha_{global}$ across observations plus effects at the country level $\alpha_{country}$ and the alert level $\alpha_{alert}$ as deviations from the global level. For the alert level effects we use Gaussian priors with mean zero and half normal standard deviations, and we use a half normal standard deviation for σ (Table X).

Other models use a similar structure, where a global effect of delay plus deviations at different levels represent the slope (Supp Tab. 4).

#### Procedure to avoid undefined ratios

In order to ensure that observed ratios would not be undefined in cases where the number of cases in the evaluation period for observations at 0 week delay is 0 (ratio denominator) and to avoid cases where the number of cases in the ratio numerator is not 0 (log(0) is undefined), we added a small constant equal to 1 to both the numerator and the denominator for all ratios, so that the log-ratio $r_i = log(\frac{y_{d_i} + 1}{y_{delay\ at\ week\ 0} + 1})$.

**Supp. Tab. 4: Equations for models to estimate the effect of additional weeks of implementation delay on the potential impact of an intervention.** Model 0 only uses a slope

$\beta$ which is assumed to be the same for all alerts. The other models use a global level slope ( $\alpha_{global}$) plus deviations at different levels ($\alpha_{country}$, $\alpha_{admin1}$, $\alpha_{loc}$, $\alpha_{alert}$ ) with half-normal hyperpriors representing their standard deviation ($\sigma_{country}$, $\sigma_{admin1}$, $\sigma_{loc}$, $\sigma_{alert}$). Measurement variability is represented by $\sigma$ .

| Model | Equations |
|---|---|
| **0** | $r_i = \log\left(\frac{y_{d_i}}{y_{\text{delay at week 0}}}\right), \quad r_i \sim N(\mu_i, \sigma), \quad \sigma \sim \text{HalfNormal}(1)$<br>$\mu_i = \beta\, d_i, \quad \beta \sim N(0,1)$ |
| **Alert** | $r_i = \log\left(\frac{y_{d_i}}{y_{\text{delay at week 0}}}\right), \quad r_i \sim N(\mu_i, \sigma), \quad \sigma \sim \text{HalfNormal}(1)$<br>$\mu_i = \beta_{\text{alert}[i]}\, d_i, \quad \beta_{\text{alert}[i]} = \alpha_{\text{global}} + \alpha_{\text{alert}[i]}$<br>$\alpha_{\text{global}} \sim N(0, \sigma_{\text{global}}), \quad \sigma_{\text{global}} \sim \text{HalfNormal}(1)$<br>$\alpha_{\text{alert}} \sim N(0, \sigma_{\text{alert}}), \quad \sigma_{\text{alert}} \sim \text{HalfNormal}(0.5)$ |
| **Country** | $r_i = \log\left(\frac{y_{d_i}}{y_{\text{delay at week 0}}}\right), \quad r_i \sim N(\mu_i, \sigma), \quad \sigma \sim \text{HalfNormal}(1)$<br>$\mu_i = \beta_{\text{country}[i]}\, d_i, \quad \beta_{\text{country}[i]} = \alpha_{\text{global}} + \alpha_{\text{country}[i]}$<br>$\alpha_{\text{global}} \sim N(0, \sigma_{\text{global}}), \quad \sigma_{\text{global}} \sim \text{HalfNormal}(1)$<br>$\alpha_{\text{country}} \sim N(0, \sigma_{\text{country}}), \quad \sigma_{\text{country}} \sim \text{HalfNormal}(1)$ |
| **Country Alert** | $r_i = \log\left(\frac{y_{d_i}}{y_{\text{delay at week 0}}}\right), \quad r_i \sim N(\mu_i, \sigma), \quad \sigma \sim \text{HalfNormal}(1)$<br>$\mu_i = \beta_{\text{alert}[i]}\, d_i, \quad \beta_{\text{alert}[i]} = \alpha_{\text{global}} + \alpha_{\text{country}[i]} + \alpha_{\text{alert}[i]}$<br>$\alpha_{\text{global}} \sim N(0, \sigma_{\text{global}}), \quad \sigma_{\text{global}} \sim \text{HalfNormal}(1)$<br>$\alpha_{\text{country}} \sim N(0, \sigma_{\text{country}}), \quad \sigma_{\text{country}} \sim \text{HalfNormal}(0.5)$<br>$\alpha_{\text{alert}} \sim N(0, \sigma_{\text{alert}}), \quad \sigma_{\text{alert}} \sim \text{HalfNormal}(0.25)$ |
| **Country Admin1 Alert** | $r_i = \log\left(\frac{y_{d_i}}{y_{\text{delay at week 0}}}\right), \quad r_i \sim N(\mu_i, \sigma), \quad \sigma \sim \text{HalfNormal}(1)$<br>$\mu_i = \beta_{\text{alert}[i]}\, d_i, \quad \beta_{\text{alert}[i]} = \alpha_{\text{global}} + \alpha_{\text{country}[i]} + \alpha_{\text{admin1}[i]} + \alpha_{\text{alert}[i]}$<br>$\alpha_{\text{global}} \sim N(0, \sigma_{\text{global}}), \quad \sigma_{\text{global}} \sim \text{HalfNormal}(1)$<br>$\alpha_{\text{country}} \sim N(0, \sigma_{\text{country}}), \quad \sigma_{\text{country}} \sim \text{HalfNormal}(0.5)$<br>$\alpha_{\text{admin1}} \sim N(0, \sigma_{\text{admin1}}), \quad \sigma_{\text{admin1}} \sim \text{HalfNormal}(0.25)$<br>$\alpha_{\text{alert}} \sim N(0, \sigma_{\text{alert}}), \quad \sigma_{\text{alert}} \sim \text{HalfNormal}(0.25)$ |
| **Location** | $r_i = \log\left(\frac{y_{d_i}}{y_{\text{delay at week 0}}}\right), \quad r_i \sim N(\mu_i, \sigma), \quad \sigma \sim \text{HalfNormal}(1)$<br>$\mu_i = \beta_{\text{location}[i]}\, d_i, \quad \beta_{\text{location}[i]} = \alpha_{\text{global}} + \alpha_{\text{location}[i]}$<br>$\alpha_{\text{global}} \sim N(0, \sigma_{\text{global}}), \quad \sigma_{\text{global}} \sim \text{HalfNormal}(1)$<br>$\alpha_{\text{location}} \sim N(0, \sigma_{\text{location}}), \quad \sigma_{\text{location}} \sim \text{HalfNormal}(1)$ |

| Model | Equations |
| --- | --- |
| **Admin1 Alert** | $r_i = \log\left(\frac{y_{d_i}}{y_{\text{delay at week 0}}}\right), \quad r_i \sim N(\mu_i, \sigma), \quad \sigma \sim \text{HalfNormal}(1)$ <br> $\mu_i = \beta_{\text{alert}[i]}\, d_i, \quad \beta_{\text{alert}[i]} = \alpha_{\text{global}} + \alpha_{\text{admin1}[i]} + \alpha_{\text{alert}[i]}$ <br> $\alpha_{\text{global}} \sim N(0, \sigma_{\text{global}}), \quad \sigma_{\text{global}} \sim \text{HalfNormal}(1)$ <br> $\alpha_{\text{admin1}} \sim N(0, \sigma_{\text{admin1}}), \quad \sigma_{\text{admin1}} \sim \text{HalfNormal}(0.5)$ <br> $\alpha_{\text{alert}} \sim N(0, \sigma_{\text{alert}}), \quad \sigma_{\text{alert}} \sim \text{HalfNormal}(0.25)$ |

### Selection of best-fit delay models

The best model for each setting was selected by examining convergence diagnostics, assessing model fit using posterior predictive checks, as well as formal model selection in order to penalize model complexity. For the model selection procedure, we compared models with different combinations of effects using Leave One Out Cross Validation (LOO) and the Watanabe Akaike Information Criterion (WAIC) using the loo package (reference). We show here an example of the model selection procedure for the ≥ 50 weekly cases for three consecutive weeks alert for the population group with 50,000 to 500,000 people.

Overall, the four models that included a global slope and alert-level deviations had the highest LOO and WAIC and were roughly equivalent given that their difference is smaller than the respective standard errors (Models B2, B3, A2, and D in the tables) (Supp Tab. 5-6). The model with the highest LOO value at -3039.18 was B2, the model with a global slope, and country-level plus alert-level deviations. Model B3, which included effects for country, administrative level 1, and alerts, had the second highest LOO value of -3039.28. Given that the standard error for those values is 52.17 and 52.22, and their difference is smaller than those values, the model fits were considered roughly equivalent. Model B2 was preferred because it has the smallest number of effective parameters (p_loo) compared to the other models with alert-level effects, suggesting greater parsimony. WAIC values reflect a similar picture. Model B2 had the highest WAIC value at -3028.70, and model B3 had the second highest WAIC value at -3039.23. Again, the standard error for those values was higher than the difference between them and model B2 had a smaller number of effective parameters (p_waic) than model B3, so model B2 was preferred.

**Supp. Tab. 5: Model comparison metrics using leave-one-out (LOO) cross-validation.**

Model Comparison using LOO for outbreak-prone locations

| Model | elpd_diff | se_diff | elpd_loo | se_elpd_loo | p_loo | se_p_loo | looic | se_looic |
|---|---|---|---|---|---|---|---|---|
| D: Global slope, admin1, and alert-specific deviation | 0.00 | 0.00 | -10070.80 | 75.85 | 227.66 | 6.12 | 20141.60 | 151.70 |
| B2: Global slope, country-specific deviation, and alert-level deviation | -0.23 | 0.69 | -10071.03 | 75.85 | 227.31 | 6.10 | 20142.06 | 151.70 |
| B3: Global slope, country, admin1, and alert-level deviation | -0.84 | 0.63 | -10071.64 | 75.85 | 227.99 | 6.13 | 20143.28 | 151.69 |
| A2: Global slope and alert-specific deviation | -1.10 | 0.63 | -10071.91 | 75.89 | 228.63 | 6.15 | 20143.81 | 151.78 |
| C: Global slope and location-specific deviation | -984.01 | 51.74 | -11054.81 | 73.24 | 197.42 | 5.28 | 22109.62 | 146.48 |
| B: Global slope and country-specific deviation | -3087.95 | 76.20 | -13158.75 | 65.60 | 18.73 | 0.67 | 26317.50 | 131.20 |
| A1: Simple null model with global rate | -3741.48 | 68.84 | -13812.29 | 52.63 | 2.18 | 0.07 | 27624.57 | 105.27 |

**Supp. Tab. 6: Model comparison metrics using the Watanabe-Akaike Information Criterion (WAIC).**

Model Comparison using WAIC for outbreak-prone locations

| Model | elpd_diff | se_diff | elpd_waic | se_elpd_waic | p_waic | se_p_waic | waic | se_waic |
|---|---|---|---|---|---|---|---|---|
| D: Global slope, admin1, and alert-specific deviation | 0.00 | 0.00 | -10069.19 | 75.83 | 226.05 | 6.06 | 20138.38 | 151.66 |
| B2: Global slope, country-specific deviation, and alert-level deviation | -0.20 | 0.68 | -10069.39 | 75.83 | 225.67 | 6.04 | 20138.78 | 151.67 |
| B3: Global slope, country, admin1, and alert-level deviation | -0.77 | 0.62 | -10069.96 | 75.83 | 226.31 | 6.07 | 20139.92 | 151.65 |
| A2: Global slope and alert-specific deviation | -1.06 | 0.63 | -10070.26 | 75.87 | 226.98 | 6.08 | 20140.51 | 151.74 |
| C: Global slope and location-specific deviation | -984.51 | 51.74 | -11053.70 | 73.23 | 196.31 | 5.24 | 22107.41 | 146.46 |
| B: Global slope and country-specific deviation | -3089.54 | 76.18 | -13158.73 | 65.60 | 18.72 | 0.67 | 26317.47 | 131.20 |
| A1: Simple null model with global rate | -3743.09 | 68.82 | -13812.29 | 52.63 | 2.18 | 0.07 | 27624.57 | 105.27 |
