## Supplemental Results for "A decision analytic framework for triggering cholera outbreak response based on early-case surveillance"

### Supplementary Results

#### Pooled location analyses

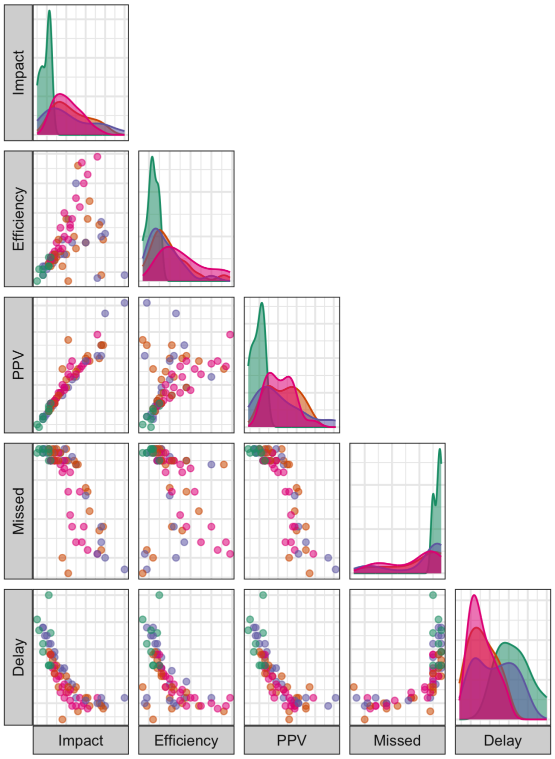


**Supp. Fig. 10. Trade-offs in utility scores and alert definition stringency across all population groups.** Panels in the bottom-left triangle are a pairs plot of the dimension-specific utility scores, where each point in the scatterplot represents a combination of alert definition and population group (24 alert definitions x 3 population groups). The distributions in the diagonal of the pairs plot show the distribution of dimension-specific utility scores. Colors represent the four alert types across all panels in the figure.

**Supp. Tab. 7 Spearman correlation coefficients between dimensions across all alert definitions and population groups.**

| Dimension1 | Dimension2 | Correlation | P_value |
|---|---|---|---|
| Impact | Efficiency | 0.89 | 0.00e+00 |
| Impact | PPV | 0.69 | 0.00e+00 |
| Impact | Missed | 0.89 | 0.00e+00 |
| Impact | Delay | -0.79 | 0.00e+00 |
| Efficiency | PPV | 0.65 | 0.00e+00 |
| Efficiency | Missed | 0.97 | 0.00e+00 |
| Efficiency | Delay | -0.80 | 0.00e+00 |
| PPV | Missed | 0.58 | 1.00e-07 |
| PPV | Delay | -0.49 | 1.06e-05 |
| Missed | Delay | -0.85 | 0.00e+00 |

#### Alert definitions enhanced with case confirmation

Increasing confirmed case thresholds were positively associated with the impact, efficiency, and PPV metrics for lower stringency weekly case-based and cumulative case-based alerts (e.g., ≥ 2 weekly cases and ≥ 5, ≥ 10, and ≥ 25 total cumulative cases for three consecutive weeks) (Supp Fig. 8). For example, the mean impact of the joint definition "≥ 2 weekly cases for three consecutive weeks and 10 confirmed cases during the alert period" was higher than the ≥ 50 weekly cases for three consecutive weeks alert definition with 0 confirmed cases (Supp Fig. 8-9). There was no apparent effect on these dimensions for higher stringency alert definitions (e.g., ≥ 100 cumulative cases over three weeks), but this may be due to the limited number of high stringency alerts triggered. By contrast, increasing confirmed case thresholds almost always led to a higher proportion of missed outbreaks and less timely alert triggers. Functionally, increasing confirmed case thresholds on top of the case-based alert definitions had a similar effect to increasing the alert definition stringency, although time series with more confirmed case data is required to fully evaluate the effect of this approach.

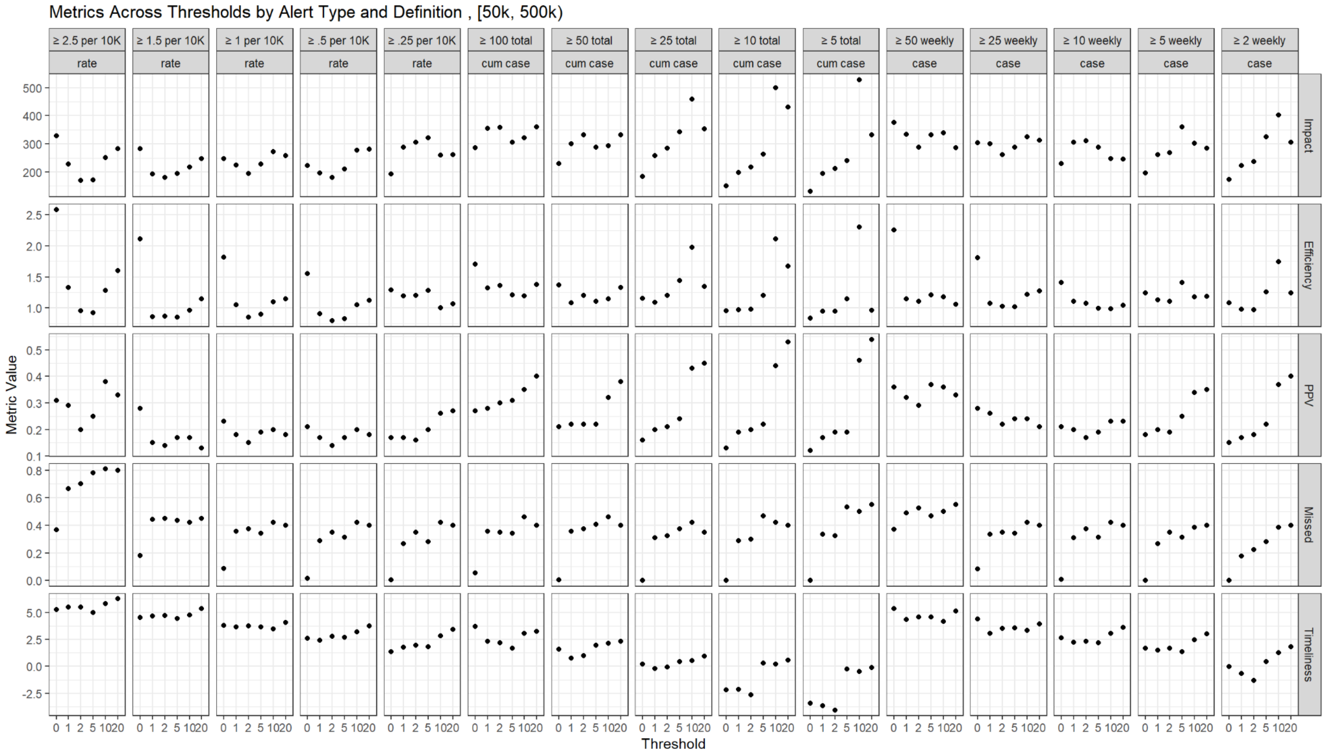


**Supp. Fig. 11: Scatterplot of mean dimension metrics (y axis) and case confirmation threshold value (x axis) for selected alert definitions of the case-based, cumulative case-based, and rate-based types triggered in locations with 50,000 to 500,000 people.**

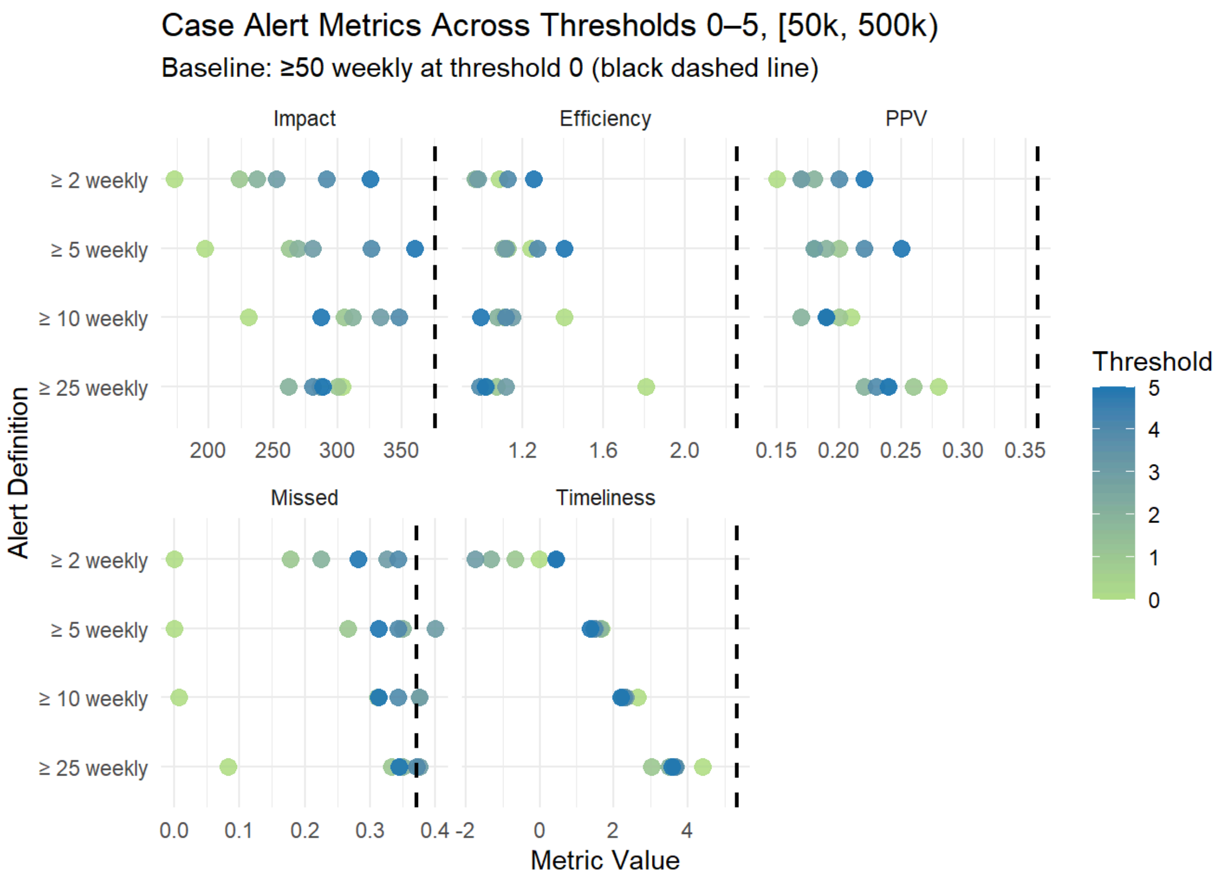


**Supp. Fig. 12. Dimension metric value (x axis) for selected alert definitions (y axis) of the case-based type for populations from 50,000 to 500,000 people compared to the dimension metric value for the ≥ 50 weekly for three consecutive weeks alert definition at case confirmation threshold 0 (no filtering), shown as a dashed vertical line.** Color reflects the case confirmation threshold.

### Country-stratified analyses

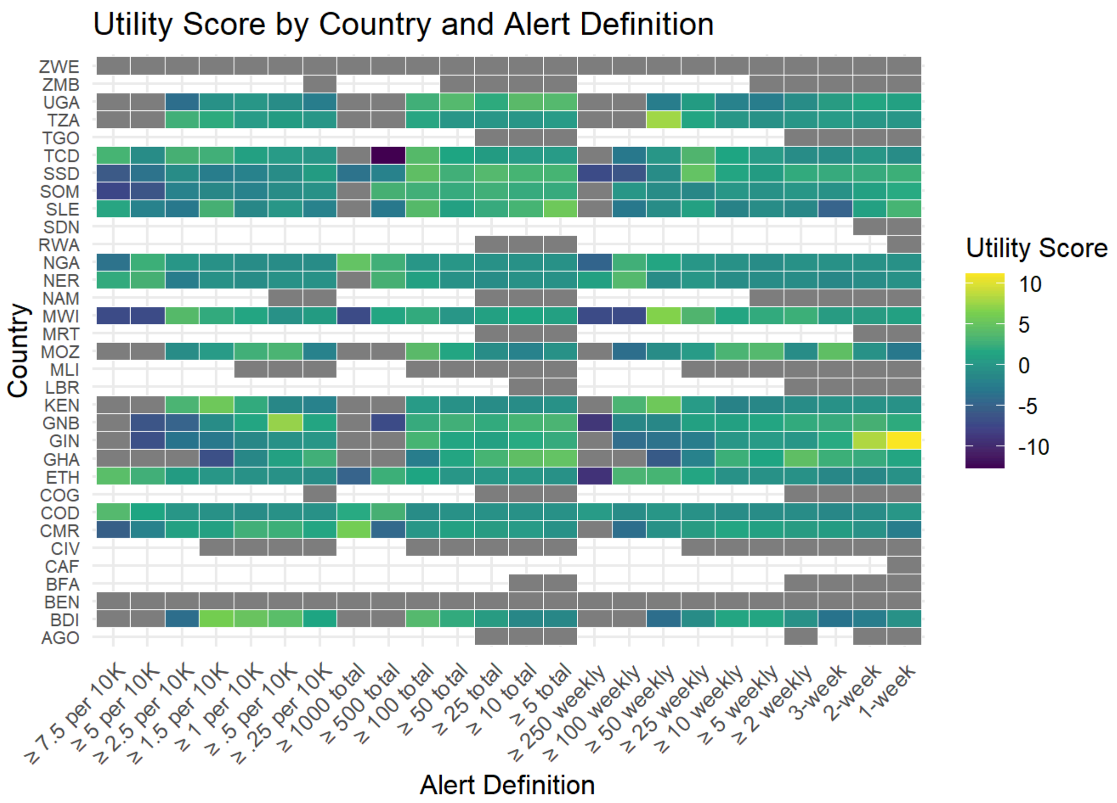


**Supp Fig 13. Heatmap of utility scores for every alert definition (on the x axis) and country (on the y axis) combination.** Color represents the utility score. Missing values for country/alert definitions are due to the fact that there were either no suspected cases in the evaluation period following these alerts in these countries, or no outbreaks linked to the alert definition/country combination.

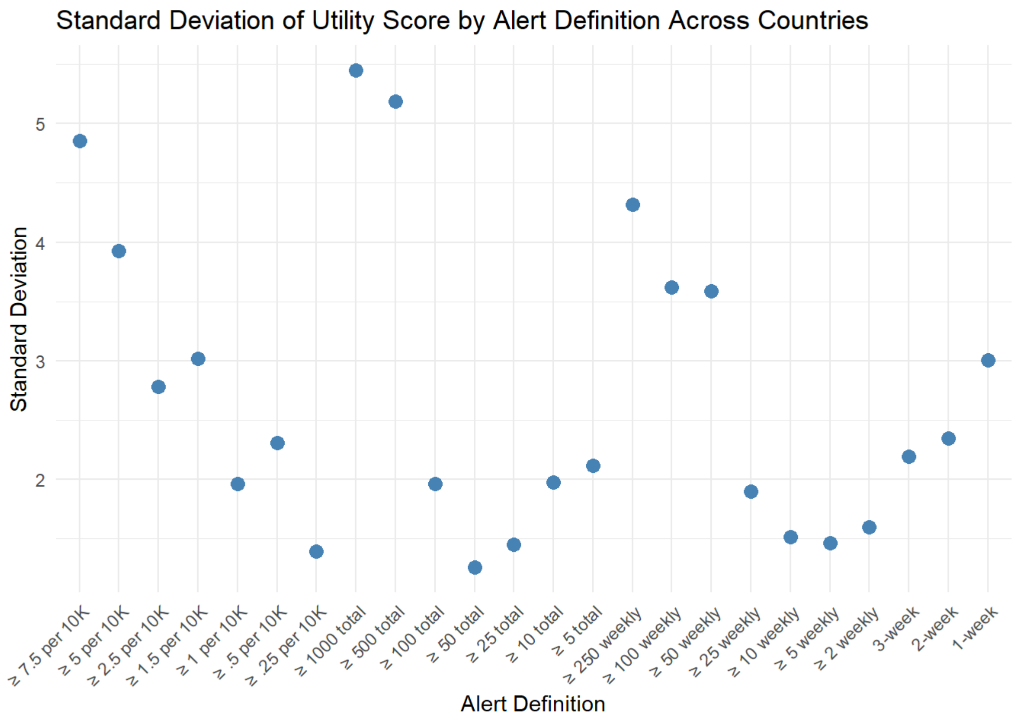


**Supp. Fig. 14. Standard deviation of utility score for every alert definition across countries.**

### Delay Model Estimates

Model with country and alert-level deviations, locations with 50,000 to 500,000 people, alert: ≥ 50 cases for 3 consecutive weeks

**Supp. Tab. 8: Posterior distribution estimates, number of effective samples, and the R-hat for the main parameters in the model fit to data from outbreak-prone locations with under 50,000 people, which incorporates country and alert-level effects as deviations from the global average slope.**

| | Posterior Estimates | | | | | | | |
|---|---|---|---|---|---|---|---|---|
| Parameter | Mean | SE_Mean | SD | 2.5% | Median | 97.5% | n_eff | Rhat |
| Global average | -0.222 | 0.001 | 0.046 | -0.320 | -0.220 | -0.136 | 3330.862 | 1.001 |
| Standard deviation of country level | 0.078 | 0.001 | 0.052 | 0.007 | 0.069 | 0.207 | 1277.315 | 1.004 |
| Standard deviation of alert level | 0.123 | 0.000 | 0.019 | 0.093 | 0.121 | 0.166 | 4716.748 | 1.001 |
| Standard deviation of measurement variability | 1.254 | 0.000 | 0.034 | 1.189 | 1.254 | 1.322 | 7989.187 | 1.000 |

**Supp. Tab. 9: Posterior distribution estimates, number of effective samples, and the R-hat for the main parameters in the model fit to data from outbreak-prone locations with 50,000 to 500,000 people, which incorporates country and alert-level effects as deviations from the global average slope.**

| | Posterior Estimates | | | | | | | |
|---|---|---|---|---|---|---|---|---|
| Parameter | Mean | SE_Mean | SD | 2.5% | Median | 97.5% | n_eff | Rhat |
| Global average | -0.221 | 0.001 | 0.019 | -0.260 | -0.220 | -0.185 | 1155.394 | 1.003 |
| Standard deviation of country level | 0.062 | 0.001 | 0.018 | 0.035 | 0.059 | 0.105 | 1273.888 | 1.001 |
| Standard deviation of alert level | 0.111 | 0.000 | 0.005 | 0.102 | 0.111 | 0.122 | 7613.113 | 1.000 |
| Standard deviation of measurement variability | 1.157 | 0.000 | 0.011 | 1.136 | 1.156 | 1.178 | 12863.783 | 1.000 |

**Supp. Tab. 10: Posterior distribution estimates, number of effective samples, and the R-hat for the main parameters in the model fit to data from outbreak-prone locations with**

**over 500,000 people, which incorporates country and alert-level effects as deviations from the global average slope.**

| | Posterior Estimates | | | | | | | |
|---|---|---|---|---|---|---|---|---|
| **Parameter** | **Mean** | **SE_Mean** | **SD** | **2.5%** | **Median** | **97.5%** | **n_eff** | **Rhat** |
| Global average | -0.222 | 0.001 | 0.046 | -0.320 | -0.220 | -0.136 | 3330.862 | 1.001 |
| Standard deviation of country level | 0.078 | 0.001 | 0.052 | 0.007 | 0.069 | 0.207 | 1277.315 | 1.004 |
| Standard deviation of alert level | 0.123 | 0.000 | 0.019 | 0.093 | 0.121 | 0.166 | 4716.748 | 1.001 |
| Standard deviation of measurement variability | 1.254 | 0.000 | 0.034 | 1.189 | 1.254 | 1.322 | 7989.187 | 1.000 |

#### Posterior Predictive Checks

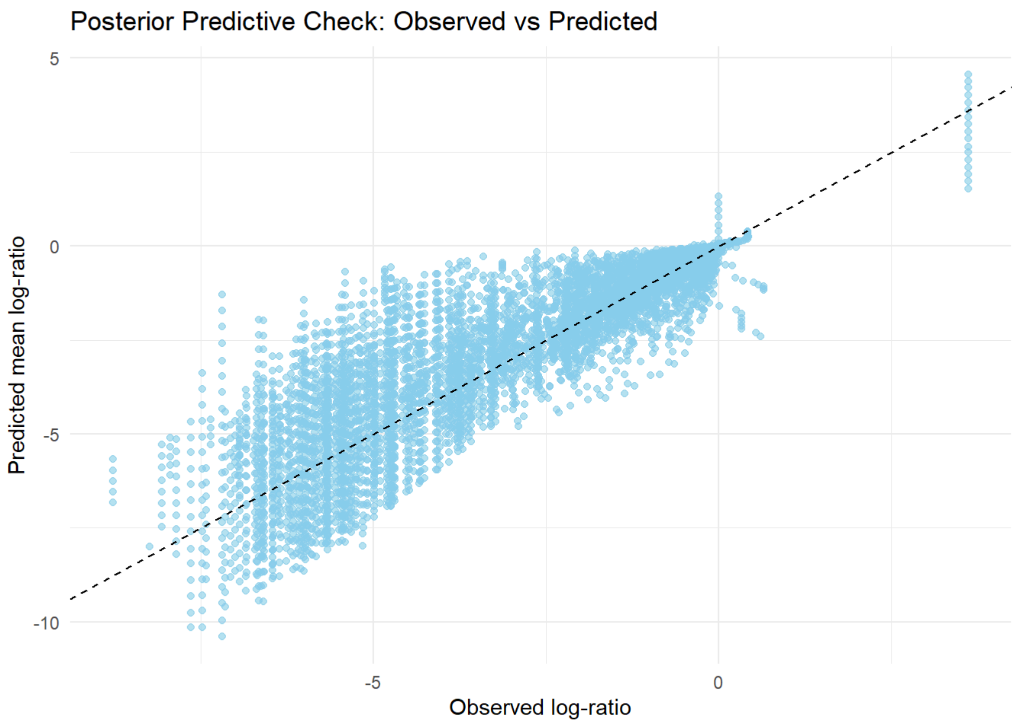


**Supp. Fig. 15: Posterior predictive check using a scatterplot with the observed log-ratio on the x axis and the mean of the posterior predicted log-ratios for that observation on the y axis for the model run for the ≥ 50 weekly cases for 3 consecutive weeks alert (with**

**global, country-level, and alert level effects) and the population group with 50,000 to 500,000 people.**

#### Country Posterior Distributions

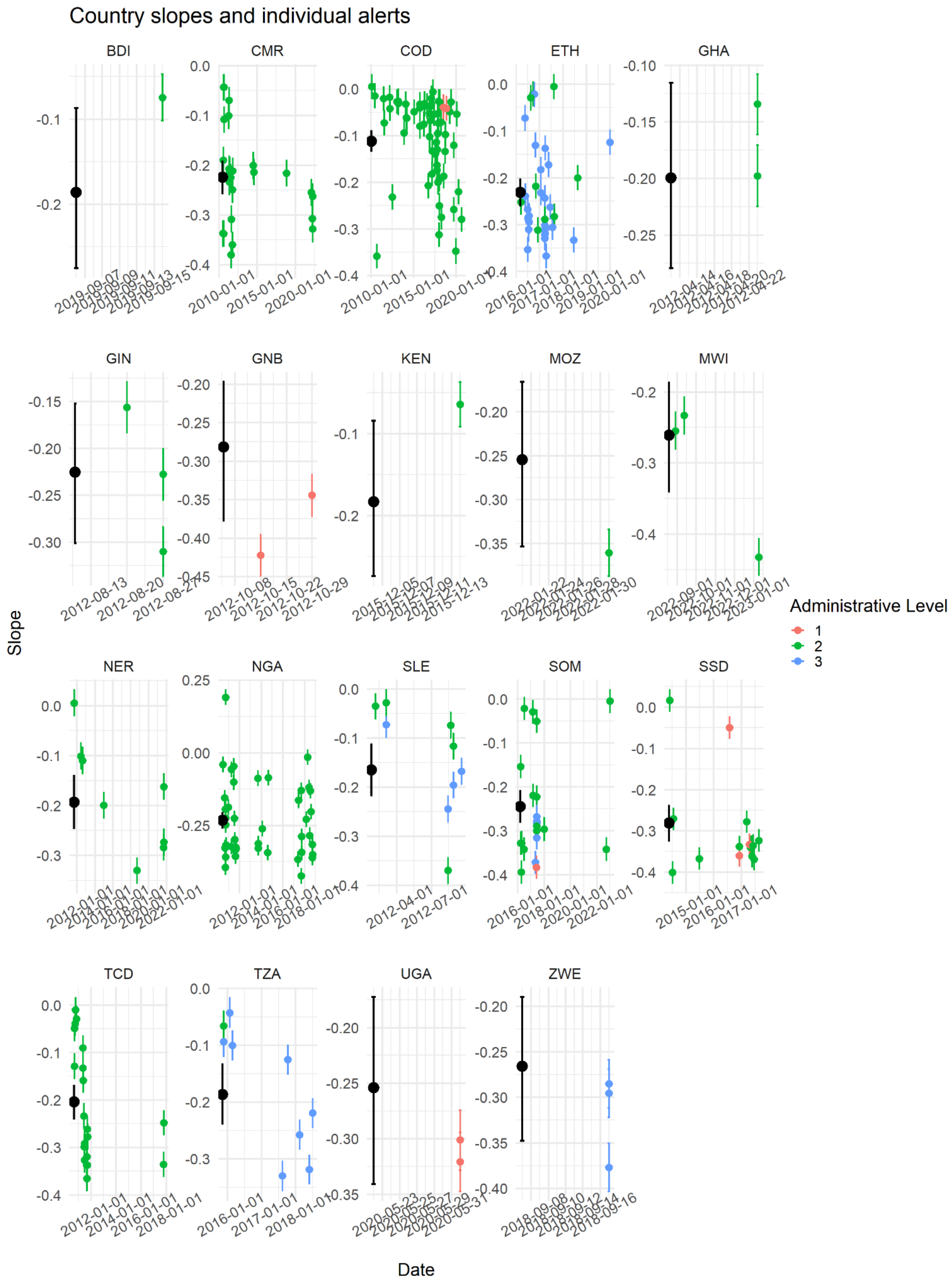

**Supp. Fig. 16: Effective slope posterior distributions (sum of global posterior and lower level deviation posterior distributions) for all alerts per country colored by administrative level, and the country-level effective slope posterior distribution in black for the alert ≥ 50 weekly cases for 3 consecutive weeks and the population group with 50,000 to 500,000 people.** For each posterior distribution we show the mean (dot) and 95 % credible interval on the y axis. The x axis shows the date when each alert was triggered.
